# Transmission lineage dynamics and the detection of viral importation in emerging epidemics

**DOI:** 10.1101/2025.03.05.25323408

**Authors:** Joseph L.-H. Tsui, Prathyush Sambaturu, Rosario Evans Pena, Linus Too, Bernardo Gutierrez, Rhys Inward, Moritz U. G. Kraemer, Louis du Plessis, Oliver G. Pybus

**Author notes:** Corresponding authors (J.L.-H.T.), (O.G.P.). M.U.G.K., L.d.P., and O.G.P. jointly supervised this work. **Competing interests:** The authors declare no competing interest.

## Abstract

The accurate inference of pathogen movements between locations during an epidemic is crucial for measuring infectious disease spread and for informing effective control strategies. Phylogeographic methods can reconstruct historical patterns of disease dissemination by combining the evolutionary history of sampled pathogen genomes with geographic information. Despite a substantial expansion of pathogen genomics during and since the COVID-19 pandemic, only a small fraction of infections are typically sampled and sequenced, leading to an underestimation of the true intensity of viral importation. Here, we seek to understand the sampling processes underlying this underestimation. We show that the coupling of viral importation and local transmission dynamics can result in local transmission lineages with different size distributions, influencing the probability that individual viral importation events will be detected. Using analytical and simulation approaches, we demonstrate that both the proportion of importation events detected and the temporal patterns of inferred importation are highly sensitive to importation dynamics and local transmission parameters. Our findings highlight the importance of interpreting phylogeographic estimates in the context of outbreak conditions, particularly when comparing viral movements across time and among different epidemic settings. These insights are critical for improving the reliability of genomic epidemiology approaches in the design of public health responses.

## 1. Introduction

Reconstruction of the spatiotemporal spread of an emerging pathogen is needed to inform the design of public health policies that aim to contain and delay further disease spread. Typically, transmission is either measured directly, for example through contact-tracing (Adam et al., 2020; Diarra et al., 2023; Faye et al., 2015; Sun et al., 2021; Vazquez-Prokopec et al., 2017) and traveller screening (Díaz-Menéndez et al., 2023; Duvignaud et al., 2024; Moritz U. G. Kraemer et al., 2020; Kucharski et al., 2023; Mayer et al., 2023; Septfons et al., 2016), or inferred indirectly from epidemiological (e.g., case incidence, hospitalisation rates) and mobility (e.g., mobile devices, flight records) data using spatial transmission models (Eggo et al., 2011; Gog et al., 2014; Viboud et al., 2006). For instance, during the 2009 H1N1 influenza pandemic, simulation models that combined intra-country commuting flows, inter-country air traffic, and high-resolution demographic data were used to characterise the dynamics and drivers of global virus spread (Bajardi et al., 2011; Balcan et al., 2009). More recently, an analysis of contact-tracing records for >600,000 COVID-19 cases in New York City during 2020-21 revealed substantial spatial heterogeneities and strong community structures, with frequent non-local transmission events across administrative regions (Pei et al., 2022). Insights from such studies can identify locations and spatial scales at which targeted interventions will be most effective and assess the potential impacts of interventions that restrict human movement.

However, epidemiological data are often constrained by reporting delays, underreporting, and logistical challenges in data collection and sharing, particularly during large outbreaks. The limited availability of real-time data on human mobility and contact patterns also hinders the accurate inference of pathogen movements, especially at small spatial scales where fine-scale heterogeneity is difficult to capture using standard mobility models (Camargo et al., 2019; Masucci et al., 2012; Yan et al., 2017). To address these limitations, pathogen genomic data are increasingly being used to investigate pathogen dissemination, with the potential to uncover cryptic transmission pathways that are not readily observed using traditional epidemiological data alone. Recent advances in high-performance computing, together with the growing availability of genomic data from public repositories such as GISAID, GenBank and Pathoplexus, have enabled the analysis of large-scale genomic and epidemiological datasets (Hill et al., 2021). These analyses often reveal complex transmission dynamics across multiple spatial scales, from transmission events among individual households to between-country movements via the global air traffic network. For example, an analysis of 482 SARS-CoV-2 sequences from students and staff at a university in the United Kingdom found limited viral introductions from the wider community, with onward transmission within the university driven primarily by shared student accommodation and in-person course-related interactions (Aggarwal et al., 2022). At the international-level, a recent study of ∼6,000 influenza genomes showed that travel and movement restrictions during the COVID-19 pandemic led to notable shifts in the global dispersal patterns of seasonal influenza lineages, with persistent transmission in South Asia during the pandemic (Chen et al., 2024).

Viral genomic epidemiology studies often rely on phylogeographic methods, which integrate information about the evolutionary relationships among pathogen genomes and the locations of sampled infections. Historical patterns of pathogen migration are inferred by extrapolating the locations of sampled sequences backward in time, guided by the ancestral relationships among them. Depending on the spatial resolution of the data and model assumptions, locations can be treated as either continuous (Lemey et al., 2010) or discrete (Lemey et al., 2009). Discrete locations are often used to model pathogens spreading among human populations due to heterogeneities in movement resulting from administrative boundaries and long-range transportation systems that span multiple spatial scales (leading to non-linearity between travel time and displacement) (Alessandretti et al., 2020; González et al., 2008; M. U. G. Kraemer et al., 2020). In discrete phylogeography models, the location of internal nodes in a phylogenetic tree (representing the ancestors of sampled viruses) are commonly inferred by modelling pathogen movement among locations as a continuous-time Markov process along an estimated phylogeny (Lemey et al., 2009). Once the internal nodes are assigned their inferred locations, local transmission clusters/lineages can be identified, with each representing a series of local transmission events that occurred in a recipient location following a single pathogen importation event. The detection and enumeration of these local transmission lineages (and their associated importation events) provides an opportunity to assess the frequency and dynamics of pathogen movement among locations.

Although analyses of local transmission lineages have been present in the literature for some time (e.g., (Hué et al., 2005), (Baillie et al., 2012)), their popularity and scale expanded during the COVID-19 pandemic, particularly in settings where traditional travel or contact tracing data were scarce or incomplete. These studies examined how viral importation from different countries contributed to the establishment of new variants and assessed the effectiveness of non-pharmaceutical interventions designed to prevent or delay further dissemination, such as travel restrictions and airport screening (Gu et al., 2022; Han et al., 2022; McCrone et al., 2021; Tsui et al., 2023). Estimates of the intensity of viral importation through time at a given location also revealed how local transmission dynamics were influenced by the introduction of new transmission lineages, with implications for the design of local control strategies. However, despite widespread adoption of these approaches, the underlying sampling processes that underpin their inferences remain poorly characterised. For instance, du Plessis et al. (du Plessis et al., 2021) investigated the early establishment of SARS-CoV-2 in the UK and found that local transmission lineages varied widely in size and were distributed heterogeneously across space and time – yet, the conditions necessary for the identification and enumeration of these lineages under such heterogeneities have not been explored. Specifically, given that only a small fraction of local infections are typically sampled and sequenced during an outbreak, to what extent is the number of viral importation events underestimated for a genomic sample of a given size, and how does this discrepancy vary over time and across different outbreak conditions?

In this study, we address this question by considering the mechanisms that underlie the detection of local transmission lineages and their associated viral importation events through discrete phylogeographic reconstruction. We show analytically how variation in viral importation intensity through time can lead to transmission lineages with different size distributions, despite the same local transmission conditions. Using a simple deterministic model, we then verify these analytical results and further show how different lineage size distributions can result in different lineage detection probabilities. Additionally, using simulated data from a stochastic agent-based model, we demonstrate the impact of temporal variation in local transmission intensity on lineage detection, and the resulting bias in the inferred importation intensity over time. We conclude that estimates of viral movements from phylogeographic analyses in the regime of low-intensity sampling () should be interpreted cautiously, and that further work is needed to mitigate such biases with consideration of both the underlying importation dynamics and local transmission conditions.

## 2. Phylogeographic reconstruction and local transmission lineages

Discrete trait analysis (DTA) (Lemey et al., 2009) has emerged as a popular approach for phylogeography due to its computational efficiency, allowing the analysis of thousands of pathogen sequences. In a typical DTA, a time-calibrated phylogeny is first estimated from a set of aligned sequences, each labelled with its location and time of sampling. The most likely location of each internal node in the estimated phylogeny is then inferred using a continuous-time Markov chain (CTMC) model. By tracing a path from the root node of the tree to each leaf node (sampled sequence), a migration or importation event is inferred to have occurred whenever we observe a change in the label going from one node to another. The inferred age of the ancestral node in a local transmission lineage (often referred to as the Time of Most Recent Common Ancestor, or TMRCA) also provides an estimate of the time of earliest detectable transmission event within that local lineage given the sampled sequences, and therefore an upper bound (most recent estimate) for the timing of viral importation (Fig. 1B) (du Plessis et al., 2021).

**Figure 1.**
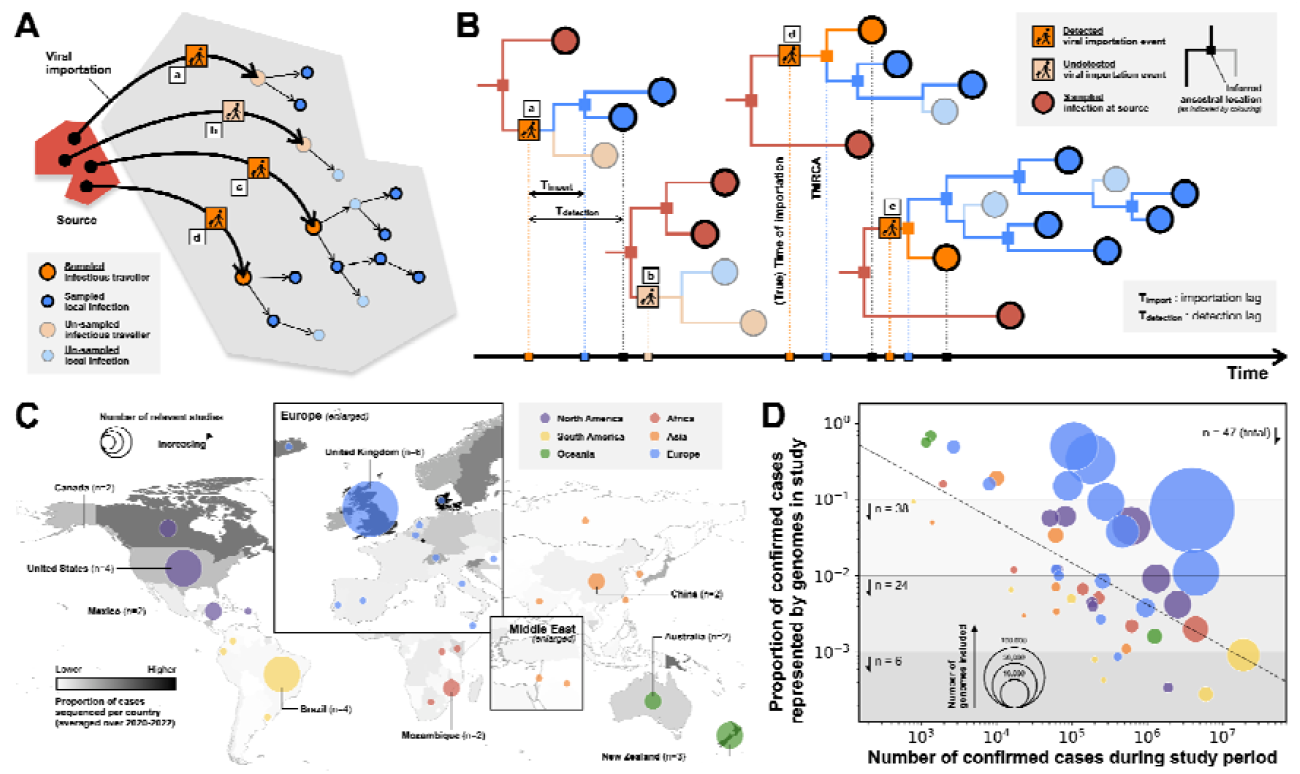
Phylogeographic reconstruction of viral importation and distribution of sampling proportions in COVID-19 studies. (A) Each viral importation event, i.e. the movement of an infectious traveller (represented as a curved arrow) from the source (red polygon) to the recipient (grey polygon) location, can be uniquely mapped to a local transmission lineage. The transmission tree associated with each local cluster is shown, with arrows representing transmission events; orange and blue circles represent arriving infectious travellers and local infections, respectively. (B) Time-scaled phylogeographic reconstruction of the local transmission lineages shown in panel (A). Internal and leaf nodes are represented as squares and circles, respectively, and coloured according to their state (red: infected and detected at source location, orange: imported and detected locally, blue: locally infected and detected). Despite complete sampling of viral genomes at the source location (red circles), only importation events (a), (c), and (d) are detected, as no members of transmission lineage (b) are sampled. The difference between the true time of importation and the TMRCA of a local transmission lineage is known as the importation lag, denoted by T_import_; the difference between the true time of importation and the time when a local transmission lineage is first detected is known as the detection lag, denoted by T_detection_. (C) Sampling proportions in previous COVID-19 studies that estimated the number of viral introductions and intensity of viral importation using phylogeographic reconstruction. Countries are coloured according to the average proportion of COVID-19 cases that were sequenced between 2020 and 2022 (darker colours indicate higher sequencing proportions). Countries from which SARS-CoV-2 genomes were collected and analysed are indicated by circles; circle radii are proportional to the number of studies for each country and coloured by continent. (D) Plot of the proportion of confirmed COVID-19 cases that were sequenced versus the total number of confirmed COVID-19 cases for each relevant study. The radius of each circle indicates the number of SARS-CoV-2 genomes included in the phylogeographic analysis of the relevant study, with colouring indicating the continent of the country in question. The black dashed line represents the least squares regression fit (Pearson’s r = - 0.63) between the log10-transformed number of confirmed cases and the log10-transformed sampling proportion. Note that a study considering four island countries across three continents was omitted from the figure; see Table S1 for more details.

Given a phylogenetic tree with internal nodes that are annotated with their most likely location, we can then partition the phylogeny into distinct, non-overlapping local transmission lineages. The use of local transmission lineages to represent a chain of transmission within a given location following a single importation event is well established in the literature on phylogenetic epidemiology (Baillie et al., 2012; Dudas et al., 2018; Hué et al., 2005) and was recently popularised by du Plessis et al. (du Plessis et al., 2021) and other studies (da Silva Filipe et al., 2021; Komissarov et al., 2021; Lemey et al., 2021) which leveraged the increased availability of viral genomic data during the COVID-19 pandemic. This formulation enables a one-to-one mapping between local transmission lineages and viral importation events (Fig. 1A, B), provided that (i) virus genetic diversity accumulates at a sufficiently rapid rate, and (ii) viral genomes at the source location (from which the pathogens are imported) are sufficiently densely sampled, such that onward local transmission lineages from distinct imported pathogen carriers can be uniquely resolved phylogenetically. In this study, we assume that both conditions are satisfied and focus our attention instead on the potential biases introduced by the undersampling of local infections.

Here we define a local transmission lineage as a group of individuals who were infected in a given location as a result of onward transmission from a single arriving infectious traveller, with the inclusion of the traveller itself. Additionally, we assume that (i) the infectious traveller remains at the recipient location indefinitely following arrival, and (ii) individuals infected locally do not travel to a new location. These assumptions together imply that:

1. Each local transmission lineage can be uniquely mapped to a single arriving infectious traveller, and therefore a single importation event.
2. Each local transmission lineage has size *l*= *n* + 1, where *n* is the total number of secondary infections that occurred locally as a result of onward transmission from the arriving infectious traveller.
3. Each local transmission lineage has a minimum size of 1, i.e. when the arriving infectious traveller fails to establish local onward transmission (i.e. *n*= 0).

Finally, we ignore any uncertainties and biases associated with phylogenetic tree estimation and ancestral state reconstruction (see discussion in Conclusion). Consequently, we can further assume that (i) there is a non-zero probability that each lineage (and its associated viral importation event) is detected by random sampling of local infections, and (ii) the sampling of at least one member of a given local transmission lineage is a sufficient and necessary condition for the detection of the lineage in a phylogeographic reconstruction.

## 3 Detection of local transmission lineages in the regime of low-intensity local sampling

Despite increases in genomic sequencing worldwide since the beginning of the COVID-19 pandemic, the number of sequenced SARS-CoV-2 genomes remains low compared to the number of reported cases in most countries. Brito et al. (Brito et al., 2022) showed that, among 189 countries with active genomic surveillance between 2020 and 2022, the average number of genomic sequences available per confirmed case is just 0.016, with only 13 countries having a sequencing coverage > 5% and 89 countries having sequencing coverage < 0.5%. We reviewed 48 phylogeographic studies that estimated the number of viral introductions at a given location (see SI. 1 and Table S1 for review details) and found that in only 9 studies did pathogen genome sequence coverage exceed 10% of the confirmed cases during the corresponding study period; in 24 studies, included sequences represented < 1% of the confirmed cases (Fig. 1B, right). We also observe a negative association between genomic sampling proportion and the number of confirmed cases, likely due to limited sequencing capacity during periods of high case incidence (Fig. 1C, D). Importantly, the true proportion of infections that were sequenced is likely lower than these estimates due to the presence of asymptomatic (Ma et al., 2021; Sah et al., 2021; Subramanian et al., 2021) and limited testing capacities, as demonstrated by the ratio of seroprevalence to cumulative incidence in many low- and middle-income countries (LMICs) (Bergeri et al., 2022; Havers et al., 2020).

Given that genome sequencing occurs at such low intensities, the number of transmission lineages detected from a sample of local infections is likely to substantially underestimate the true number of extant transmission lineages in the population. More formally, given *S* genomic samples collected at random from a local population of infections of size *N*_*I*_, the probability of detecting a lineage of size *l* within the sample is given by

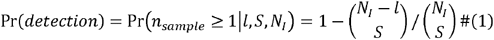

where *n*_*sample*_ is the number of lineage members that are sampled. In the regime of low-intensity sampling such that *S*≪ *N*_*l*_, we obtain the approximation

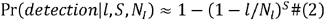

Using this approximation, the proportion of lineages in an infected population that we can expect to find in a sample of size *s* (referred to as lineage detection probability *r*_*d*_ hereafter) is therefore given by

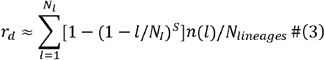

where *n*(*l*), hereafter referred to as lineage density at *l*, is the number of lineages of size *l* in the local infected population, such that 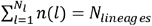, the total number of lineages in the local infected population. Note that the summation has an upper bound *N*_*l*_, corresponding to the maximum size of a lineage in the scenario where there is only a single transmission lineage.

From the above result, we can see that the lineage detection probability depends on not only the sample size *S* (relative to the size of the infected population *N*_*l*_) (Fig. 2B), but also the lineage size distribution, as specified by *n* (*l*). We can draw useful insights by considering a hypothetical scenario in which each lineage is of size *l*= 1 (i.e. when none of the arriving infectious travellers are able to establish local onward transmission), for which we obtain *r*_*d*_ ≈ *S*/*N*_*I*_ from Eq. 3, i.e. the expected number of detected lineages is directly proportional to the sampling proportion, *s*= *s*/*N*_*I*_. Conversely, in the hypothetical scenario where the entire infected population consists of a single transmission lineage of size *N*_*I*_, Eq. 3 reduces to *r*_*d*_ =1 for all *S*> 0 and *l* ≥ 1, i.e. only a single sampled infection is needed to detect all importation events (with *N*_*lineages*_ = 1), as expected. In reality, the underlying lineage size distribution, and therefore the expected behaviour of lineage detection, will lie somewhere between these two extremes, as we show below.

**Figure 2.**
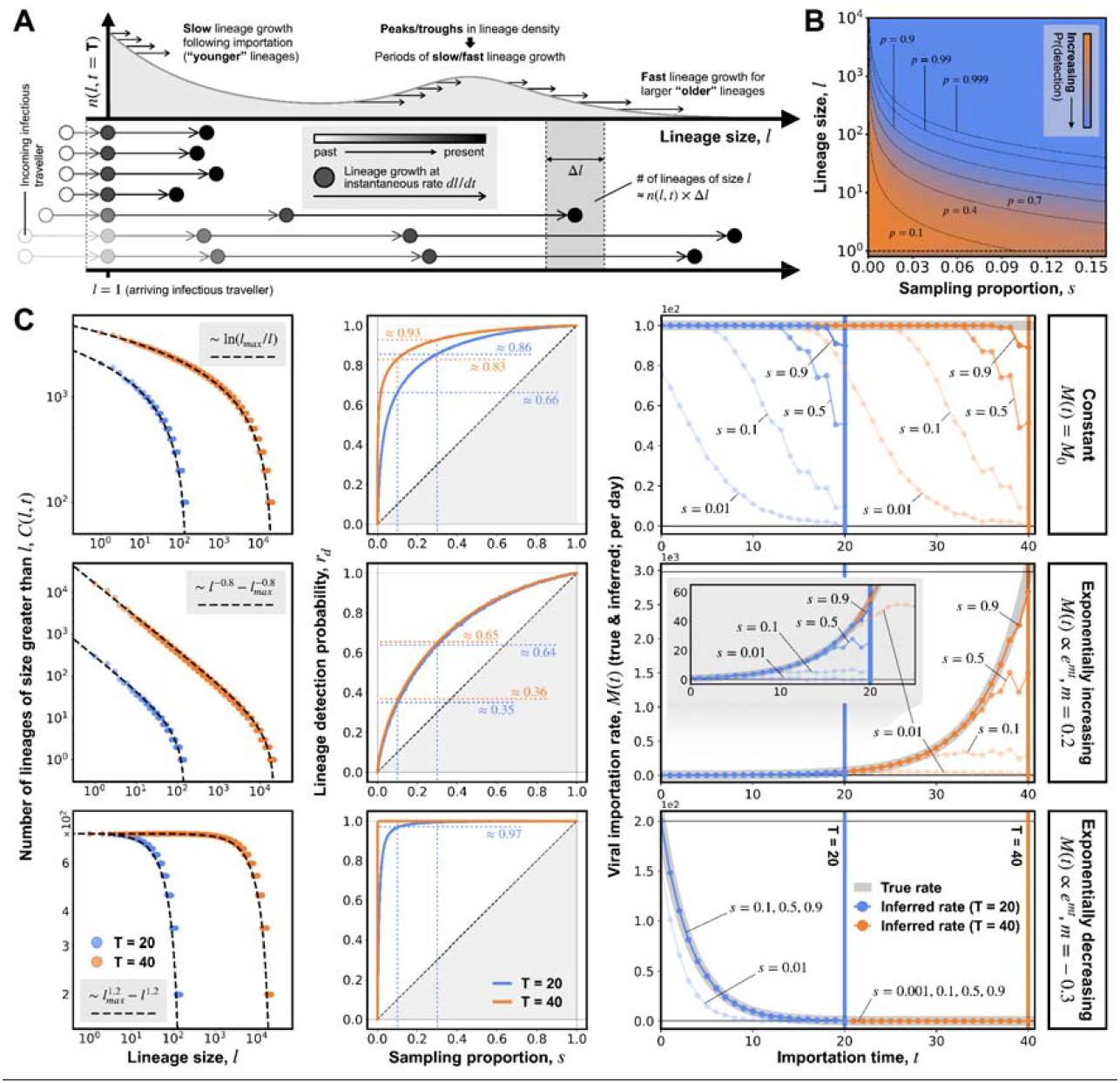
Time evolution of local transmission lineage size distribution and simulated lineage detection under a simple deterministic model with local exponential growth. (A) An illustration of the time evolution of the transmission lineage size distribution in an outbreak modelled as the movement of particles in a 1D lineage size-space. Each particle represents a single local transmission lineage, with instantaneous velocity in size-space given by its growth rate dl/dt, which we assume to be homogeneous across lineages of the same size at a given time. Variation in local growth rate as a function of lineage size leads to different size distributions, e.g., slower growth rate of smaller lineages tends to result in a lineage size distribution that decays with increasing lineage size. (B) Probability of detecting a local transmission lineage from a random sample of local infections (total number of infections N_I_ = 10^6^), as a function of lineage size l (y-axis) and sampling proportion S (x-axis). (C) Results from simulated lineage detection under a simple deterministic model assuming no recovery process and no depletion of susceptibles, with local exponential growth at rate r = 0.25. Each row presents results from simulations under a different scenario of importation dynamics. The three scenarios (see labelled boxes on the far-right of each row) are: a constant importation rate (top row), an exponentially increasing importation rate (middle row) and an exponentially decreasing importation rate (bottom row). See figure and text for parameter details. The left hand column compares the cumulative frequency distributions of lineage sizes from simulations (coloured circles) with those obtained from analytical solutions (dashed black lines). Results are shown for two different observation times (blue: T=20; orange: T=40). The middle column shows the median proportion of lineages detected (or lineage detection probability r_d_) at different sampling proportions. Results are shown for two different observation times (blue: T=20; orange: T=40). The right hand column compares the true importation rate (solid grey line) with the inferred importation rate (dotted lines) at different sampling proportions. Results are again shown for two different observation times; vertical lines indicate the times when lineage detection is simulated (blue: T=20; orange: T=40). The inset on row 2 enlarges the same data in the interval 0≤t≤ 25.

Several studies have attempted to measure the size distribution of transmission lineages in an infected population, with most of which reporting observed distributions that were right-skewed (heavy-tailed) (du Plessis et al., 2021; Murall et al., 2021; Tsui et al., 2023), i.e. many small lineages and only a few large ones. In practice, however, the true underlying size distribution of local transmission lineages in an infected population cannot be measured directly, especially in the regime of low-intensity sampling, because the observed size of a detected lineage depends on both the sampling proportion and the true lineage size, with the latter being an unknown quantity (see Fig. S1). This challenge closely resembles the ecological problem of estimating the number of unseen species, where the observed relative species abundance distribution is skewed by incomplete sampling with rare species being underrepresented (Chao and Lee, 1992; Efron and Thisted, 1976; Fisher et al., 1943; Hubbell, 2011; Volkov et al., 2003). We leave this non-trivial inference problem for future work. Instead, we use a simple analytical model to investigate how different lineage size distributions may arise as a result of the coupling between viral importation and local transmission.

## 4 Time evolution of transmission lineage size distribution

We consider a hypothetical scenario in which the number of infectious travellers arriving at a location of interest from an arbitrary source location is given by *M*(*t*) for *t*> 0. In particular, we focus our attention on the early phase of an epidemic or the emergence of a new immune escape variant of a known pathogen, when it can be reasonably assumed that (i) the population at the location of interest (referred to as the local population hereafter) is completely susceptible, (ii) the rate of local transmission is sufficiently high such that the recovery of infectious individuals has a negligible impact on the overall dynamics of the local outbreak, and (iii) there is no substantial depletion of susceptibles. We will later relax assumptions (ii) and (iii) in a simulation analysis.

Given these assumptions, and following from the abovementioned definition of a local transmission lineage, we propose that the growth of a transmission lineage following the arrival of an infectious traveller can be modelled as the movement of a particle in a one-dimensional continuous lineage size-space (given a sufficiently large infected population) (Fig. 2A). The time evolution of the density of these particles (each representing a single local transmission lineage) in this size-space can therefore be considered as solutions to the continuity equation

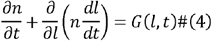

with the boundary condition *n* (*l*,0) = 0 for all *l* ≥ 1, where *n*= *n* (*l)* is the particle density at *l* in size-space, i.e. the number of lineages of size between *l* and *l*+ Δ*l*, and is analogous to the variable with the same notation in Eq. 3, in the limit of large *l. dl*/*dt* represents the instantaneous lineage growth rate, which we assume to be homogeneous across lineages of the same size at a given time. In the fluid dynamics literature (Kundu et al., 2012; White, 2000), the term *G* (*l, t*) is commonly referred to as a *source term*, which here describes the rate at which new transmission lineages are being introduced to the location of interest through importation. Assuming further that there is no outward movement of infected individuals, and since each transmission lineage must have a minimum size of *l*= 1 (as described previously), we set

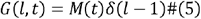

where *δ* (*l* − 1) is the Dirac delta function centred at *l*= 1.

Depending on the assumed functional form of the lineage growth rate and importation rate, the above partial differential equation (Eq. 4) can be solved either analytically or numerically (e.g., using finite difference methods). Here we consider a hypothetical scenario in which the local infected population is experiencing exponential growth such that *dl*/*dt* = *rl*, where *r* is a positive constant, as is commonly assumed during the early stages of an outbreak. It can be shown that Eq. 4 can be solved analytically to give the solution (see SI. 2)

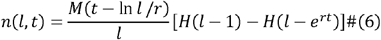

where *H* (*x*) is the Heaviside step function. The term *H* (*l* − 1) *H* (*l −e*^*rT*^) is commonly known as a boxcar function, and is zero everywhere except for the interval *l* ∈ [1,*e*^*rT*^]in which it takes a value of 1. The right-limit of this interval, *e*^*rT*^, corresponds to the maximum size attainable for a lineage seeded at *t*= 0 under the assumption of local exponential growth and therefore the maximum possible lineage size at a given time *t* since the first importation event; similarly, the left-limit of this interval represents the minimum size of a lineage (i.e. *l*= 1).

Importantly, we have not made any assumptions in our derivation regarding the functional form of the underlying importation rate, *M*(*t*) It is therefore instructive to consider the behaviour of *n* (*l,t*) under different assumptions of *M* (*t*), specifically (i) a constant rate, (ii) an exponentially decreasing rate, and (iii) an exponentially increasing rate:

1. Assuming a constant rate of viral importation (*M* (*t*)= *M*_0_), the first term in Eq. 6 reduces to *M*_0_/*l*, i.e. the lineage density is inversely proportional to lineage size *l*. This result is not surprising, as larger lineages grow more rapidly compared to smaller lineages under the assumption of exponential growth. In our model of lineage growth, this implies that particles corresponding to larger lineages move at a higher velocity towards the right, resulting in a lower lineage density at large (Fig. 2A).
2. Conversely, the lower velocity of particles corresponding to smaller lineages results in their accumulation at small ; the higher the rate at which new particles (arriving infectious travellers) are introduced relative to their velocity in size-space (instantaneous lineage growth rate), the more rapidly these particles accumulate. Indeed, if we assume an exponentially decreasing importation rate (, with), the first term in Eq. 6 reduces to. This represents a power-law distribution, which increases in density with only if, i.e. only when the rate at which new lineages are introduced is decreasing sufficiently rapidly compared to the local growth rate that we obtain lineage density that increases with lineage size.
3. During the early dissemination of an emerging pathogen, however, the rate of viral importation is likely to be increasing over time as prevalence at the origin location increases, in which case we retrieve a lineage size distribution that decreases in density with increasing lineage size following a power-law, i.e. with.

The derivation of an equivalent expression for under the assumption of local logistic growth (e.g., as a result of the depletion of susceptibles as the outbreak progresses) can be found in SI. 3.

## 5. Lineage detection under local exponential growth

Having derived an expression (Eq. 6) that describes the time evolution of the underlying lineage size distribution in an infected population experiencing local exponential growth (with no recovery process and no depletion of susceptibles), next we consider the implications of such an evolution on lineage detection. Specifically, we consider the impact of changes in the lineage size distribution on (i) the proportion of lineages and therefore viral importation events that are likely to be detected during a period of observation, and (ii) trends in the inferred importation rate over time. To do so, we construct a simple deterministic model in which new lineages of size *l*= 1 are introduced at rate *M*(*t*) Each new lineage is assumed to undergo deterministic exponential growth with no recovery of infected individuals. We run each simulation up to a predefined time T, at which point we perform random sampling of all infected individuals and identify their corresponding local transmission lineages to simulate the inference process of a phylogeographic reconstruction (see SI. 4 for details).

Using this simple model, we first verify the analytical results derived above by examining the lineage size distributions that result from different importation dynamic scenarios. Given the discrete nature of lineage size, it is preferable to compare the (inverse) cumulative lineage size distribution (denoted by *C* (*l,t*), i.e. the number of lineages of size ≥ *l* at a given time *t*) (du Plessis et al., 2021), instead of the actual lineage size distribution *n*(*l,t*). The corresponding analytical expression for C (*l,t*) under the assumption of a constant importation rate can be found by integrating Eq. 6 from *l* up to *l*_*max*_ (*T*) =*e*^*rT*^ (the maximum lineage size attainable at time of observation T), giving

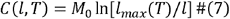

under the assumption of a constant rate of importation, and

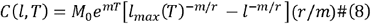

under the assumption of an exponentially increasing (*m*> 0) or decreasing (*m*< 0) rate of importation.

From Fig. 2C (left column), we see that the simulated lineage size distributions are in good agreement with the analytical predictions, with some deviations at small lineage sizes, likely due to the continuous approximation of discrete lineage size used in our analytical derivation (see SI. 2). Interestingly, in the case of an exponentially decreasing importation rate, we observe that *C*(*l,T*) approaches the same value in the limit *l* → 1 at both times of observation (Fig. 2C). This can be explained by noting that *C* (*l* = 1,*T*)= *N*_*lineages*_(*T*), i.e. the total number of extant lineages in the local infected population at time T. As a result of the rapidly decreasing rate of importation, the total number of lineages remains fixed between *t*= 20 and *t*= 40 while the lineages that have already been seeded continue to grow to larger sizes (as indicated by the apparent rightward shift of the observed cumulative frequency distribution, *C* (*l,T*), between *t*=20 and *t*=40). These observations together indicate a lineage size distribution that increases in density with lineage size, as expected given |*m*| > *r*(*m*= − 0.3, *r* = 0.25).

Given the different lineage size distributions that result from different (true) importation rates, it is unsurprising that we would observe different lineage detection probabilities *r*_*d*_ and, more specifically, different behaviours in *r*_*d*_ as a function of sampling proportion *s* (Fig. 2C, middle column). We find that when importation rate is exponentially decreasing, *r*_*d*_ rapidly approaches 1 at small *s* (≲ 0.01); whereas when importation is exponentially increasing, *r*_*d*_ only exceeds 50% at *s* ≈ 0.2. This observed difference can be explained by considering the lineage size distribution: the distribution resulting from an exponentially decreasing importation rate has a greater proportion of larger lineages, whereas a right-skewed distribution with a long tail is predicted for a constant or exponentially increasing importation rate. This difference is accentuated at large T, as lineages that have already been seeded continue to grow, while the total number of extant lineages approaches a finite value if importation rate is exponentially decreasing, but increases over time for a constant or exponentially increasing importation (Fig. 2C, left column).

Lineages seeded earlier are more likely to be detected at a given time of observation T given our assumption of local exponential growth, as they have more time to grow to larger sizes than recently seeded lineages. We observe this effect in Fig. 2C (right column), where the proportion of detected importation events decreases through time. Consequently the trends in inferred importation are consistently downwardly biased compared to the true importation rate, with greater discrepancy close to the time of observation. This also leads to different inferred importation trends depending on the time of observation; for example, in the case of exponentially increasing importation (Fig. 2C, right column, middle row), an early analysis at T=20 at low sampling proportions (e.g. *s* ≲ 0.1) would conclude erroneously that the intensity of viral importation has started to slow or even decrease, while a later observation and analysis at T=40 would indicate a continually increasing importation rate at T=20. Note that the extent of this discrepancy also depends on the rate of local lineage growth, with slower growth giving rise to less variation in lineage size and therefore a less pronounced reduction in inferred importation rate close to the time of observation.

## 6. Lineage detection under constant and time-varying local contact rates with recovery

We have assumed so far that the depletion of susceptibles is negligible, which is only likely to be valid during the early stages of an outbreak when the number of infected individuals is small. Further, we have assumed that (i) there is no recovery process, and (ii) each local transmission lineage grows deterministically following its introduction. Together, these two assumptions imply that lineages that are introduced earlier will always grow to larger sizes than those introduced later, regardless of local transmission dynamics. In practice, this is unlikely to be true, especially during periods of low transmission intensity (e.g., due to depletion of susceptibles or epidemic control interventions), when lineages might be subject to stochastic extinction and stop growing soon after introduction (Curran-Sebastian et al., 2025).

To explore the impact of these assumptions, we construct an agent-based model with a stochastic transmission process in which, at each time step, susceptible individuals become infected with a certain probability upon contact with an infectious individual. Individuals are assumed to mix randomly regardless of their infection status. We also assume a stochastic recovery process, by which infectious individuals recover and become immune to further infection probabilistically at a constant rate. Following the same procedure as in the previous section, we simulate the detection of viral importation events by randomly sampling infected individuals at a given time of observation (T) and identifying their corresponding local transmission lineages. Here we keep the rate of viral importation constant (100 infectious travellers arriving per day) and instead vary the local transmission dynamics. Specifically, we explore the detection of transmission lineages under two scenarios: (i) fixed local transmission conditions and (ii) time-varying local transmission conditions, in which the contact rate changes through time following a sigmoidal trajectory (see Table S2 for details).

Results of the first scenario are shown in Fig. 3. As in the deterministic model (Fig 2C), the lineage detection probability *r*_*d*_ varies with sampling proportion *s* (Fig. 3A-C). Notably, we find a slower increase of *r*_*d*_ with *s* when individuals can recover from infection, as compared to without recovery, at both T=10 (Fig. 3A) and T=30 (Fig. 3B). This is unsurprising, as the recovery of infectious individuals leads to slower lineage growth and therefore smaller lineages at the time of observation. However, this pattern is reversed at T=50 (Fig. 3C); this can be explained by the depletion of susceptibles after T=50 under the assumption of no recovery (Fig. 3D, blue line), resulting in later-introduced lineages being smaller at observation and therefore less likely to be detected. In some cases these later-introduced lineages immediately become extinct (see Fig. S2). In contrast, the rate of depletion of the susceptible population is slower when individuals can recover from infection (Fig. 3H, blue line), resulting in larger lineages at later time points.

**Figure 3.**
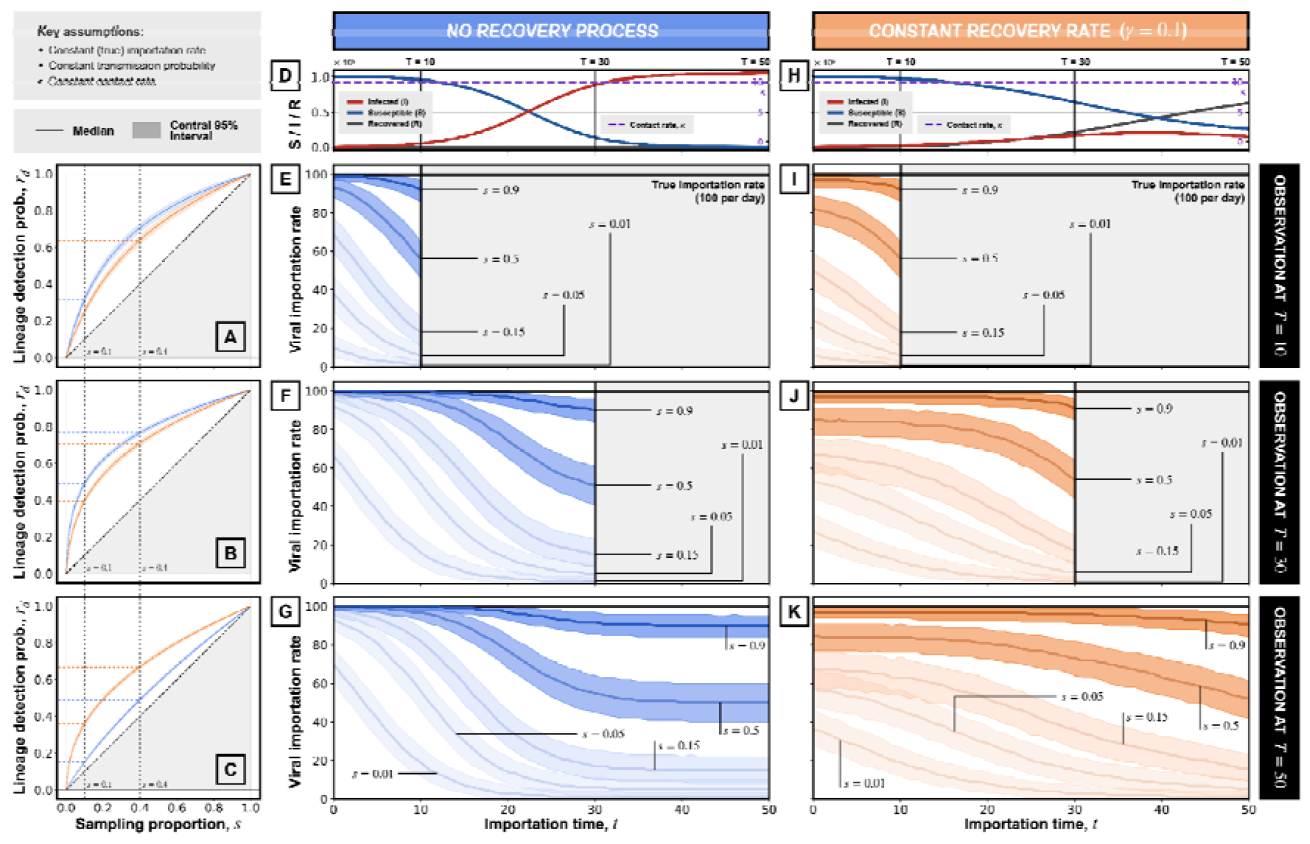
Simulated lineage detection in a stochastic agent-based model assuming a constant local contact rate. (D, H) Simulated epidemic dynamics, showing the number of infected (I; red solid line), susceptible (S; blue solid line) and recovered (R; black solid line) individuals through time, averaged across 200 simulation replicates. Simulation parameters: importation rate per day; transmission probability per contact; contact rate per day. The left column (A-C) shows the proportion of lineages detected for different sampling proportions (across simulation replicates). Solid lines and shaded regions represent the median and the central 95% interval, respectively, of the proportion of detected lineages (with blue and orange indicating results from simulations assuming no recovery process and a constant recovery rate, respectively). The middle (E-G) and right (I-K) columns show the inferred importation rate through time (across simulation replicates), assuming no recovery process (blue) and a constant recovery rate (orange), respectively. Results are shown for different sampling proportions, s (shading transparency varies with s). Solid lines and shaded regions show the median and central 95% interval, respectively, of the inferred importation rate. Solid black lines represent the true importation rate (100 per day). Each row shows results for a different observation time, as indicated by labels on the far-right (T=10, 30, 50).

This effect is also apparent in the inferred importation rates, especially at T=50 under the assumption of no recovery (Fig. 3G), for which we observe a sharp drop in the inferred rate at as a result of rapid depletion of susceptibles, resulting in later-introduced lineages being smaller and thus less likely to be detected. Conversely, in the case of a constant recovery rate, we observe a more gradual decline in the inferred importation rate (Fig. 3K).

Fig. 4 shows results for the second simulated scenario, in which the contact rate varies over time. We observe substantial differences in the lineage detection probability between simulations in which increases versus those in which decreases. In the case of increasing, there is an almost linear relationship between and at T=10 (Fig. 4A, blue line), i.e. when is changing most rapidly (Fig. 4D, purple dashed line). This can be explained by a substantially lower contact rate before T=10 which gives rise to slower lineage growth and therefore more frequent lineage extinction and smaller lineages at T=10; consequently the lineage detection probability scales almost linearly with sampling proportion, as described in Eq. 2. This effect is also reflected in the observation that the inferred importation rate barely changes through time when is increasing (Fig. 4E), whereas the inferred rate abruptly declines into the recent past when is decreasing (Fig. 4I). Again, this is because the higher contact rate before results in earlier-introduced lineages being larger and therefore more likely to be detected compared to later-introduced lineages.

**Figure 4.**
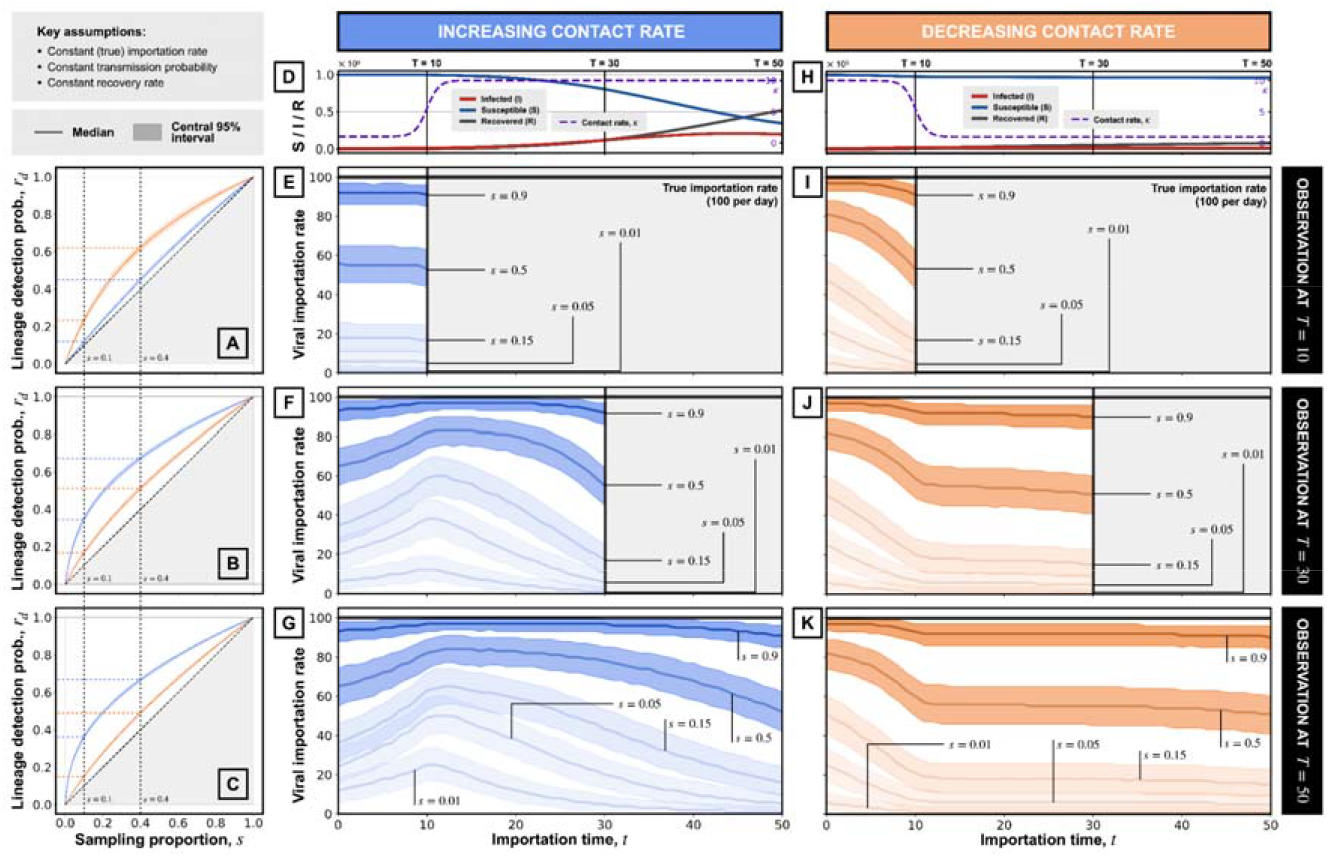
Simulated lineage detection in a stochastic agent-based model assuming time-varying contact rates. (D, H) Simulated epidemic dynamics, showing the number of infected (I; red solid line), susceptible (S; blue solid line) and recovered (R; black solid line) individuals through time, averaged across 200 simulation replicates. Simulation parameters: importation rate per day; transmission probability per contact; recovery rate per day. The left column (A-C) shows the proportion of lineages detected for different sampling proportions (across simulation replicates). Solid lines and shaded regions represent the median and the central 95% interval, respectively, of the proportion of detected lineages (with blue and orange indicating results from simulations assuming no recovery process and a constant recovery rate, respectively). The middle (E-G) and right (I-K) columns show the inferred importation rate through time (across simulation replicates), assuming an increasing (blue) and decreasing (orange) local contact rate, respectively. Results are shown for different sampling proportions, s (shading transparency varies with s). Solid lines and shaded regions show the median and central 95% interval, respectively, of the inferred importation rate. Solid black lines represent the true importation rate (100 per day). Each row shows results for a different observation time, as indicated by labels on the far-right (T=10, 30, 50).

At T=30 and T=50, these observed patterns are reversed, as expected given the abrupt changes in at T=10. Notably, we see that the lineage detection probability increases more slowly with in the case of decreasing (Fig. 4B, C), as a result of new lineages either growing very slowly or becoming extinct soon after introduction. Interestingly, in the case of increasing, the higher contact rate following T=10 leads to an apparent increase in the inferred importation rate initially (Fig. 4F, G), as later-introduced lineages are more likely to survive past T=10 beyond which they can continue to grow at higher. This is then followed by a rapid decline towards the time of observation, again due to variation in lineage size resulting from different importation times; whereas in the case of decreasing, we observe a slower decline (Fig. 4J, K) due to smaller size variation among lineages introduced at small after T=10. Importantly, these abrupt changes in the inferred rate lead to mischaracterisation of the underlying trend of viral importation - despite the true importation rate being constant through time, the degree to which it is underestimated varies substantially over time as a function of both the time-varying local transmission dynamics (the contact rate in the simulations) and sampling proportion *s*.

## Conclusion

Phylogeographic analyses have enabled the detection and enumeration of viral importation events, necessary for understanding the dispersal patterns of an emerging pathogen and evaluating the impact of public health interventions, especially those designed to limit further spatial dissemination. Standard phylogeographic approaches, such as discrete trait analysis, rely on the identification of local transmission lineages consisting of local secondary infections that result from arriving infectious travellers. Given that only a small fraction of local infections are typically sequenced in an outbreak, it is unsurprising that the number of viral importation events inferred from a given genomic dataset underestimates the true number. However, the underlying mechanism that leads to this underestimation and the degree to which this occurs under different outbreak conditions has not been well characterised.

Here, we showed that the proportion of viral importation events detected from a sample of local infections depends on the underlying lineage size distribution, which in turn is determined by the coupled dynamics of viral importation and local transmission. By modelling lineage growth as particle movements in a continuous size-space, we found that the lineage size distribution of an infected population undergoing local exponential growth can be described by a power-law (consistent with empirical observations of SARS-CoV-2 spread made by du Plessis et al. (du Plessis et al., 2021) and other similar studies (McCrone et al., 2021; Murall et al., 2021; Tsui et al., 2023)), with the exponent depending on both local lineage growth rate and viral importation intensity over time. More generally, when local lineage growth is slower than the rate at which new lineages are being introduced, there is an accumulation of smaller lineages and, as a result, the proportion of lineages that we expect to detect increases only slowly with sample size. In contrast, when local transmission is intense, lineages grow rapidly compared to the importation rate, resulting in larger lineages that are more likely to be detected, even at low sampling proportions.

Using an agent-based stochastic model, we found that the inferred importation rate can be substantially downwardly biased, especially at low sampling intensities. This is due to variation in the lineage size distribution at the time of observation, which itself is determined by (i) differences in the time elapsed since introduction, with recently-introduced lineages being smaller and therefore harder to detect, and (ii) stochastic extinction of local lineages during periods of low transmission, potentially resulting in smaller lineages at the time of observation despite early importation.

Our findings have implications for the interpretation of viral movement estimates from phylogeographic analyses. First, while it is known that these estimates represent only approximate lower bounds of the true number of viral importation events, our results indicate that the degree of underestimation can vary substantially between outbreak contexts, especially when the fraction of infections that are genomically sequenced is small. This is important in the context of studies that investigate the source-sink dynamics of virus spread, in which the relative intensities of viral movement between locations are of primary interest. Our findings suggest that such estimates could be substantially biased and their interpretation requires careful consideration of the differences in local transmission dynamics between locations, especially when sampling fractions are low (≲ 5%). Second, our observation that inferred viral importation dynamics can depend on local transmission intensity has implications for the interpretation of such estimates in the context of policy evaluation. For instance, a phylogeographic analysis of virus genomes from a country that experienced a recent increase in transmission intensity might conclude erroneously that the resurgence in case numbers was driven by increased viral importation - in reality, the true importation intensity might have remained constant, with the apparent increase being an artefact resulting from later-introduced lineages being larger and therefore more likely to be detected (see Fig. 4F, G). Conversely, an increase in viral importation intensity shortly before the time of genome sampling is unlikely to be detected, due to the lower detection probability of recently-introduced (and therefore smaller) lineages. Studies that evaluate the efficacy of public health policies, especially those intended to prevent or reduce viral importation, should therefore be cautious in interpreting the temporal dynamics of viral movement estimates from phylogeographic analyses when sampling intensities are low.

There are several limitations to our approach. Throughout the study we assumed sufficient sampling of virus genetic diversity at the source location, such that each independent local lineage can be uniquely resolved phylogenetically. This assumption rarely holds in practice due to both limited sequencing capacity and under-reporting, which will apply equally to both the source and recipient location. Violation of this assumption leads to an aggregation of independent local lineages and therefore an underestimation of the true number of viral importation events, even in the limit of complete sampling of infected individuals at the recipient location. Future studies should explore the extent to which the aggregation of lineages occurs given different sampling proportions at the source location and the effect of variation in relevant epidemiological and evolutionary parameters, as well as potential mitigation strategies when presented with such sampling bias (Lemey et al., 2020). Secondly, we also assumed that each infected individual at the recipient location is equally likely to be sampled. This may not hold in practice due to targeted or biased sampling across space (e.g., unequal access to testing or healthcare services, airport screening, intense sampling following contact-tracing) or time (e.g., changes in public awareness, testing fatigue, sequencing capacity being overwhelmed during intense transmission). This will likely lead to further biases in the inferred viral movements, especially if the sampling probability is correlated with viral importation rate or local transmission intensity; future studies involving empirical data should consider the potential impact of these sampling heterogeneities. Thirdly, by randomly sampling infected individuals only at the end of an outbreak, we have implicitly assumed that local transmission dynamics are unaffected by the sampling process. While this is a reasonable assumption for low-intensity sampling (the focus of this study), future work could consider different types of sampling or sequencing efforts (for example, when infected individuals are required to self-isolate following a positive test result) and their impact on transmission dynamics at high sampling intensities. Finally, we assumed that statistical uncertainties associated with phylogenetic and phylogeographic inference are negligible and can be ignored. In reality, the choice of molecular clock models (Tay et al., 2024), tree priors (Featherstone et al., 2022), and phylogeographic methods (e.g., structured coalescent (De Maio et al., 2015; Müller et al., 2017; Vaughan et al., 2014), continuous random walk (Lemey et al., 2010)) can all impact estimates of viral lineage movement. Further, we assumed that the timing of importation associated with each local transmission lineage is known - in practice, the exact time when an infectious traveller arrives can be obscured by both importation-lag (i.e. time difference between importation and first inferred local transmission event) and detection-lag (i.e. time difference between importation and first detected local infection; see Fig 1B). These lags remain poorly understood and have received little attention in the literature. A more comprehensive evaluation of our findings in the context of these uncertainties should be explored.

As large-scale pathogen genomic sequencing becomes more common in global public health, phylogeographic analysis is likely to play an increasingly important role in reconstructing and monitoring the spread of emerging pathogens. While this study primarily focuses on characterising the sampling process underlying the detection of viral importation events, our findings lay the groundwork for developing more robust inference methods to address these biases. Such methods are needed to derive more accurate insights from viral genomic data and to inform the design of effective, timely interventions in response to future epidemics.

## Data Availability

All data, code and analysis files used in this work are openly available from GitHub at https://github.com/joetsui1994/Transmission-lineage-dynamics-and-detection/.

## Data, Materials, and Software Availability

### Acknowledgements

M.U.G.K. acknowledges funding from The Rockefeller Foundation (PC-2022-POP-005), Google.org, the Oxford Martin School Programmes in Pandemic Genomics (also O.G.P., J.L.-H.T.) & Digital Pandemic Preparedness, European Union’s Horizon Europe programme projects MOOD (#874850) and E4Warning (#101086640), Wellcome Trust grants 303666/Z/23/Z, 226052/Z/22/Z & 228186/Z/23/Z, the United Kingdom Research and Innovation (#APP8583), the Medical Research Foundation (MRF-RG-ICCH-2022-100069), UK International Development (301542-403), the Bill & Melinda Gates Foundation (INV-063472) and Novo Nordisk Foundation (NNF24OC0094346). J.L.-H.T. is supported by a Yeotown Scholarship from New College, University of Oxford. The contents of this publication are the sole responsibility of the authors and do not necessarily reflect the views of the European Commission or the other funders.

## Author contributions

J.L.-H.T. designed the research. J.L.-H.T., R.E.P. performed the research and analysed data. L.d.P., P.S., L.T., M.U.G.K., O.G.P. advised on methods. J.L.-H.T. wrote the initial manuscript with critical input from all authors. All authors edited and revised the manuscript.

## References

Adam, D.C., Wu, P., Wong, J.Y., Lau, E.H.Y., Tsang, T.K., Cauchemez, S., Leung, G.M., Cowling, B.J., 2020. Clustering and superspreading potential of SARS-CoV-2 infections in Hong Kong. Nature Medicine 26, 1714–1719.

Aggarwal, D., Warne, B., Jahun, A.S., Hamilton, W.L., Fieldman, T., du Plessis, L., Hill, V., Blane, B., Watkins, E., Wright, E., Hall, G., Ludden, C., Myers, R., Hosmillo, M., Chaudhry, Y., Pinckert, M.L., Georgana, I., Izuagbe, R., Leek, D., Nsonwu, O., Hughes, G.J., Packer, S., Page, A.J., Metaxaki, M., Fuller, S., Weale, G., Holgate, J., Brown, C.A., Howes, R., McFarlane, D., Dougan, G., Pybus, O.G., Angelis, D.D., Maxwell, P.H., Peacock, S.J., Weekes, M.P., Illingworth, C., Harrison, E.M., Matheson, N.J., Goodfellow, I.G., 2022. Genomic epidemiology of SARS-CoV-2 in a UK university identifies dynamics of transmission. Nat. Commun. 13, 1–16.

Alessandretti, L., Aslak, U., Lehmann, S., 2020. The scales of human mobility. Nature 587, 402–407.

Baillie, G.J., Galiano, M., Agapow, P.-M., Myers, R., Chiam, R., Gall, A., Palser, A.L., Watson, S.J., Hedge, J., Underwood, A., Platt, S., McLean, E., Pebody, R.G., Rambaut, A., Green, J., Daniels, R., Pybus, O.G., Kellam, P., Zambon, M., 2012. Evolutionary Dynamics of Local Pandemic H1N1/2009 Influenza Virus Lineages Revealed by Whole-Genome Analysis. J. Virol. 10.1128/jvi.05347-11

Bajardi, P., Poletto, C., Ramasco, J.J., Tizzoni, M., Colizza, V., Vespignani, A., 2011. Human Mobility Networks, Travel Restrictions, and the Global Spread of 2009 H1N1 Pandemic. PLOS ONE 6, e16591.

Balcan, D., Colizza, V., Gonçalves, B., Hu, H., Ramasco, J.J., Vespignani, A., 2009. Multiscale mobility networks and the spatial spreading of infectious diseases. Proceedings of the National Academy of Sciences 106, 21484–21489.

Bergeri, I., Whelan, M.G., Ware, H., Subissi, L., Nardone, A., Lewis, H.C., Li, Z., Ma, X., Valenciano, M., Cheng, B., Al Ariqi, L., Rashidian, A., Okeibunor, J., Azim, T., Wijesinghe, P., Le, L.-V., Vaughan, A., Pebody, R., Vicari, A., Yan, T., Yanes-Lane, M., Cao, C., Clifton, D.A., Cheng, M.P., Papenburg, J., Buckeridge, D., Bobrovitz, N., Arora, R.K., Van Kerkhove, M.D., Unity Studies Collaborator Group, 2022. Global SARS-CoV-2 seroprevalence from January 2020 to April 2022: A systematic review and meta-analysis of standardized population-based studies. PLOS Medicine 19, e1004107.

Brito, A.F., Semenova, E., Dudas, G., Hassler, G.W., Kalinich, C.C., Kraemer, M.U.G., Ho, J., Tegally, H., Githinji, G., Agoti, C.N., Matkin, L.E., Whittaker, C., Bulgarian SARS-CoV-2 sequencing group, Communicable Diseases Genomics Network (Australia and New Zealand), COVID-19 Impact Project, Danish Covid-19 Genome Consortium, Fiocruz COVID-19 Genomic Surveillance Network, GISAID core curation team, Network for Genomic Surveillance in South Africa (NGS-SA), Swiss SARS-CoV-2 Sequencing Consortium, Howden, B.P., Sintchenko, V., Zuckerman, N.S., Mor, O., Blankenship, H.M., de Oliveira, T., Lin, R.T.P., Siqueira, M.M., Resende, P.C., Vasconcelos, A.T.R., Spilki, F.R., Aguiar, R.S., Alexiev, I., Ivanov, I.N., Philipova, I., Carrington, C.V.F., Sahadeo, N.S.D., Branda, B., Gurry, C., Maurer-Stroh, S., Naidoo, D., von Eije, K.J., Perkins, M.D., van Kerkhove, M., Hill, S.C., Sabino, E.C., Pybus, O.G., Dye, C., Bhatt, S., Flaxman, S., Suchard, M.A., Grubaugh, N.D., Baele, G., Faria, N.R., 2022. Global disparities in SARS-CoV-2 genomic surveillance. Nat. Commun. 13, 7003.

Camargo, C.Q., Bright, J., Hale, S.A., 2019. Diagnosing the performance of human mobility models at small spatial scales using volunteered geographical information. Royal Society Open Science. 10.1098/rsos.191034

Chao, A., Lee, S.-M., 1992. Estimating the Number of Classes via Sample Coverage. Journal of the American Statistical Association.

Chen, Z., Tsui, J.L.-H., Gutierrez, B., Moreno, S.B., du Plessis, L., Deng, X., Cai, J., Bajaj, S., Suchard, M.A., Pybus, O.G., Lemey, P., Kraemer, M.U.G., Yu, H., 2024. COVID-19 pandemic interventions reshaped the global dispersal of seasonal influenza viruses. Science. 10.1126/science.adq3003

Curran-Sebastian, J., Andersen, F.M., Bhatt, S., 2025. Modelling the stochastic importation dynamics and establishment of novel pathogenic strains using a general branching processes framework. Math Biosci 380, 109352.

da Silva Filipe, A., Shepherd, J.G., Williams, T., Hughes, J., Aranday-Cortes, E., Asamaphan, P., Ashraf, S., Balcazar, C., Brunker, K., Campbell, A., Carmichael, S., Davis, C., Dewar, R., Gallagher, M.D., Gunson, R., Hill, V., Ho, A., Jackson, B., James, E., Jesudason, N., Johnson, N., McWilliam Leitch, E.C., Li, K., MacLean, A., Mair, D., McAllister, D.A., McCrone, J.T., McDonald, S.E., McHugh, M.P., Morris, A.K., Nichols, J., Niebel, M., Nomikou, K., Orton, R.J., O’Toole, Á., Palmarini, M., Parcell, B.J., Parr, Y.A., Rambaut, A., Rooke, S., Shaaban, S., Shah, R., Singer, J.B., Smollett, K., Starinskij, I., Tong, L., Sreenu, V.B., Wastnedge, E., COVID-19 Genomics UK (COG-UK) Consortium, Holden, M.T.G., Robertson, D.L., Templeton, K., Thomson, E.C., 2021. Genomic epidemiology reveals multiple introductions of SARS-CoV-2 from mainland Europe into Scotland. Nat Microbiol 6, 112–122.

De Maio, N., Wu, C.-H., O’Reilly, K.M., Wilson, D., 2015. New Routes to Phylogeography: A Bayesian Structured Coalescent Approximation. PLoS Genet 11, e1005421.

Diarra, M., Ndiaye, R., Barry, A., Talla, C., Diagne, M.M., Dia, N., Faye, J., Sarr, F.D., Gaye, A., Diallo, A., Cisse, M., Dieng, I., Fall, G., Tall, A., Faye, O., Faye, O., Sall, A.A., Loucoubar, C., 2023. Analysis of contact tracing data showed contribution of asymptomatic and non-severe infections to the maintenance of SARS-CoV-2 transmission in Senegal. Scientific Reports 13, 1–9.

Díaz-Menéndez, M., Angelo, K.M., de Miguel Buckley, R., Bottieau, E., Huits, R., Grobusch, M.P., Gobbi, F.G., Asgeirsson, H., Duvignaud, A., Norman, F.F., Javelle, E., Epelboin, L., Rothe, C., Chappuis, F., Martinez, G.E., Popescu, C., Camprubí-Ferrer, D., Molina, I., Odolini, S., Barkati, S., Kuhn, S., Vaughan, S., McCarthy, A., Lago, M., Libman, M.D., Hamer, D.H., 2023. Dengue outbreak amongst travellers returning from Cuba—GeoSentinel surveillance network, January–September 2022. Journal of travel medicine 30, taac139.

Dudas, G., Carvalho, L.M., Rambaut, A., Bedford, T., 2018. MERS-CoV spillover at the camel-human interface. 10.7554/eLife.31257

du Plessis, L., McCrone, J.T., Zarebski, A.E., Hill, V., Ruis, C., Gutierrez, B., Raghwani, J., Ashworth, J., Colquhoun, R., Connor, T.R., Faria, N.R., Jackson, B., Loman, N.J., O’Toole, Á., Nicholls, S.M., Parag, K.V., Scher, E., Vasylyeva, T.I., Volz, E.M., Watts, A., Bogoch, I.I., Khan, K., COVID-19 Genomics UK (COG-UK) Consortium, Aanensen, D.M., Kraemer, M.U.G., Rambaut, A., Pybus, O.G., 2021. Establishment and lineage dynamics of the SARS-CoV-2 epidemic in the UK. Science 371, 708–712.

Duvignaud, A., Stoney, R.J., Angelo, K.M., Chen, L.H., Cattaneo, P., Motta, L., Gobbi, F.G., Bottieau, E., Bourque, D.L., Popescu, C.P., Glans, H., Asgeirsson, H., Oliveira-Souto, I., Vaughan, S.D., Amatya, B., Norman, F.F., Waggoner, J., Díaz-Menéndez, M., Beadsworth, M., Odolini, S., Camprubí-Ferrer, D., Epelboin, L., Connor, B.A., Eperon, G., Schwartz, E., Libman, M., Malvy, D., Hamer, D.H., Huits, R., GeoSentinel Network, 2024. Epidemiology of travel-associated dengue from 2007 to 2022: A GeoSentinel analysis. J Travel Med 31. 10.1093/jtm/taae089

Efron, B., Thisted, R., 1976. Estimating the number of unseen species: How many words did Shakespeare know? Biometrika 63. 10.1093/BIOMET/63.3.435

Eggo, R.M., Cauchemez, S., Ferguson, N.M., 2011. Spatial dynamics of the 1918 influenza pandemic in England, Wales and the United States. Journal of The Royal Society Interface. 10.1098/rsif.2010.0216

Faye, O., Boëlle, P.-Y., Heleze, E., Faye, O., Loucoubar, C., Magassouba, N. ‘faly, Soropogui, B., Keita, S., Gakou, T., El Hadji Ibrahima, B., Koivogui, L., Sall, A.A., Cauchemez, S., 2015. Chains of transmission and control of Ebola virus disease in Conakry, Guinea, in 2014: an observational study. The Lancet Infectious Diseases 15, 320–326.

Featherstone, L.A., Zhang, J.M., Vaughan, T.G., Duchene, S., 2022. Epidemiological inference from pathogen genomes: A review of phylodynamic models and applications. Virus Evol 8, veac045.

Fisher, R., Corbet, A., Williams, C.B., 1943. The Relation Between the Number of Species and the Number of Individuals in a Random Sample of an Animal Population. Journal of Animal Ecology. 10.2307/1411

Gog, J.R., Ballesteros, S., Viboud, C., Simonsen, L., Bjornstad, O.N., Shaman, J., Chao, D.L., Khan, F., Grenfell, B.T., 2014. Spatial Transmission of 2009 Pandemic Influenza in the US. PLOS Computational Biology 10, e1003635.

González, M.C., Hidalgo, C.A., Barabási, A.-L., 2008. Understanding individual human mobility patterns. Nature 453, 779–782.

Gu, H., Xie, R., Adam, D.C., Tsui, J.L.-H., Chu, D.K., Chang, L.D.J., Cheuk, S.S.Y., Gurung, S., Krishnan, P., Ng, D.Y.M., Liu, G.Y.Z., Wan, C.K.C., Cheng, S.S.M., Edwards, K.M., Leung, K.S.M., Wu, J.T., Tsang, D.N.C., Leung, G.M., Cowling, B.J., Peiris, M., Lam, T.T.Y., Dhanasekaran, V., Poon, L.L.M., 2022. Genomic epidemiology of SARS-CoV-2 under an elimination strategy in Hong Kong. Nat. Commun. 13, 1–10.

Han, A.X., Kozanli, E., Koopsen, J., Vennema, H., RIVM COVID-19 molecular epidemiology group, Aarts, L., Bos, S., van den Brandt, A., van den Brink, S., Cremer, J., Freriks, K., Jaarsma, R., Schmitz, D., Then, E., van der Veer, B., Wijsman, L., Zwagemaker, F., Hajji, K., Kroneman, A., van Walle, I., Klinkenberg, D., Wallinga, J., Russell, C.A., Eggink, D., Reusken, C., 2022. Regional importation and asymmetric within-country spread of SARS-CoV-2 variants of concern in the Netherlands. 10.7554/eLife.78770

Havers, F.P., Reed, C., Lim, T., Montgomery, J.M., Klena, J.D., Hall, A.J., Fry, A.M., Cannon, D.L., Chiang, C.F., Gibbons, A., Krapiunaya, I., Morales-Betoulle, M., Roguski, K., Rasheed, M.A.U., Freeman, B., Lester, S., Mills, L., Carroll, D.S., Owen, S.M., Johnson, J.A., Semenova, V., Blackmore, C., Blog, D., Chai, S.J., Dunn, A., Hand, J., Jain, S., Lindquist, S., Lynfield, R., Pritchard, S., Sokol, T., Sosa, L., Turabelidze, G., Watkins, S.M., Wiesman, J., Williams, R.W., Yendell, S., Schiffer, J., Thornburg, N.J., 2020. Seroprevalence of Antibodies to SARS-CoV-2 in 10 Sites in the United States, March 23-May 12, 2020. JAMA Intern. Med. 10.1001/jamainternmed.2020.4130

Hill, V., Ruis, C., Bajaj, S., Pybus, O.G., Kraemer, M.U.G., 2021. Progress and challenges in virus genomic epidemiology. Trends in parasitology 37. 10.1016/j.pt.2021.08.007

Hubbell, S.P., 2011. The Unified Neutral Theory of Biodiversity and Biogeography. Princeton University Press.

Hué, S., Pillay, D., Clewley, J.P., Pybus, O.G., 2005. Genetic analysis reveals the complex structure of HIV-1 transmission within defined risk groups. Proc. Natl. Acad. Sci. U. S. A. 102, 4425–4429.

Komissarov, A.B., Safina, K.R., Garushyants, S.K., Fadeev, A.V., Sergeeva, M.V., Ivanova, A.A., Danilenko, D.M., Lioznov, D., Shneider, O.V., Shvyrev, N., Spirin, V., Glyzin, D., Shchur, V., Bazykin, G.A., 2021. Genomic epidemiology of the early stages of the SARS-CoV-2 outbreak in Russia. Nat. Commun. 12, 1–13.

Kraemer, M.U.G., Sadilek, A., Zhang, Q., Marchal, N.A., Tuli, G., Cohn, E.L., Hswen, Y., Perkins, T.A., Smith, D.L., Reiner, R.C., Brownstein, J.S., 2020. Mapping global variation in human mobility. Nature human behaviour 4. 10.1038/s41562-020-0875-0

Kraemer, M.U.G., Yang, C.-H., Gutierrez, B., Wu, C.-H., Klein, B., Pigott, D.M., Open COVID-19 Data Working Group†, du Plessis, L., Faria, N.R., Li, R., Hanage, W.P., Brownstein, J.S., Layan, M., Vespignani, A., Tian, H., Dye, C., Pybus, O.G., Scarpino, S.V., 2020. The effect of human mobility and control measures on the COVID-19 epidemic in China. Science. 10.1126/science.abb4218

Kucharski, A.J., Chung, K., Aubry, M., Teiti, I., Teissier, A., Richard, V., Russell, T.W., Bos, R., Olivier, S., Cao-Lormeau, V.-M., 2023. Real-time surveillance of international SARS-CoV-2 prevalence using systematic traveller arrival screening: An observational study. PLOS Medicine 20, e1004283.

Kundu, P.K., Cohen, I.M., Dowling, D.R., 2012. Fluid Mechanics. Academic Press.

Lemey, P., Hong, S.L., Hill, V., Baele, G., Poletto, C., Colizza, V., O’Toole, Á., McCrone, J.T., Andersen, K.G., Worobey, M., Nelson, M.I., Rambaut, A., Suchard, M.A., 2020. Accommodating individual travel history and unsampled diversity in Bayesian phylogeographic inference of SARS-CoV-2. Nature Communications 11, 1–14.

Lemey, P., Rambaut, A., Drummond, A.J., Suchard, M.A., 2009. Bayesian phylogeography finds its roots. PLoS computational biology 5. 10.1371/journal.pcbi.1000520

Lemey, P., Rambaut, A., Welch, J.J., Suchard, M.A., 2010. Phylogeography takes a relaxed random walk in continuous space and time. Mol. Biol. Evol. 27, 1877–1885.

Lemey, P., Ruktanonchai, N., Hong, S.L., Colizza, V., Poletto, C., Van den Broeck, F., Gill, M.S., Ji, X., Levasseur, A., Oude Munnink, B.B., Koopmans, M., Sadilek, A., Lai, S., Tatem, A.J., Baele, G., Suchard, M.A., Dellicour, S., 2021. Untangling introductions and persistence in COVID-19 resurgence in Europe. Nature 595, 713–717.

Ma, Q., Liu, J., Liu, Q., Kang, L., Liu, R., Jing, W., Wu, Y., Liu, M., 2021. Global Percentage of Asymptomatic SARS-CoV-2 Infections Among the Tested Population and Individuals With Confirmed COVID-19 Diagnosis: A Systematic Review and Meta-analysis. JAMA network open 4. 10.1001/jamanetworkopen.2021.37257

Masucci, A.P., Serras, J., Johansson, A., Batty, M., 2012. Gravity vs radiation model: on the importance of scale and heterogeneity in commuting flows. 10.1103/PhysRevE.88.022812

Mayer, A.B., Consigny, P.H., Grobusch, M.P., Camprubí-Ferrer, D., Huits, R., Rothe, C., 2023. Chikungunya in returning travellers from Bali - A GeoSentinel case series. Travel medicine and infectious disease 52. 10.1016/j.tmaid.2023.102543

McCrone, J.T., Hill, V., Bajaj, S., Pena, R.E., Lambert, B.C., Inward, R., Bhatt, S., Volz, E., Ruis, C., Dellicour, S., Baele, G., Zarebski, A.E., Sadilek, A., Wu, N., Schneider, A., Ji, X., Raghwani, J., Jackson, B., Colquhoun, R., O’Toole, Á., Peacock, T.P., Twohig, K., Thelwall, S., Dabrera, G., Myers, R., COVID-19 genomics UK (COG-UK) consortium, Faria, N.R., Huber, C., Bogoch, I.I., Khan, K., du Plessis, L., Barrett, J.C., Aanensen, D.M., Barclay, W.S., Chand, M., Connor, T., Loman, N.J., Suchard, M.A., Pybus, O.G., Rambaut, A., Kraemer, M.U.G., 2021. Context-specific emergence and growth of the SARS-CoV-2 Delta variant. medRxiv.

Müller, N.F., Rasmussen, D.A., Stadler, T., 2017. The Structured Coalescent and Its Approximations. Mol Biol Evol 34, 2970–2981.

Murall, C.L., Fournier, E., Galvez, J.H., N’Guessan, A., Reiling, S.J., Quirion, P.-O., Naderi, S., Roy, A.-M., Chen, S.-H., Stretenowich, P., Bourgey, M., Bujold, D., Gregoire, R., Lepage, P., St-Cyr, J., Willet, P., Dion, R., Charest, H., Lathrop, M., Roger, M., Bourque, G., Ragoussis, J., Shapiro, B.J., Moreira, S., 2021. A small number of early introductions seeded widespread transmission of SARS-CoV-2 in Québec, Canada. Genome Med. 13, 1–17.

Pei, S., Kandula, S., Cascante Vega, J., Yang, W., Foerster, S., Thompson, C., Baumgartner, J., Ahuja, S.D., Blaney, K., Varma, J.K., Long, T., Shaman, J., 2022. Contact tracing reveals community transmission of COVID-19 in New York City. Nature Communications 13, 1–8.

Sah, P., Fitzpatrick, M.C., Zimmer, C.F., Abdollahi, E., Juden-Kelly, L., Moghadas, S.M., Singer, B.H., Galvani, A.P., 2021. Asymptomatic SARS-CoV-2 infection: A systematic review and meta-analysis. Proceedings of the National Academy of Sciences 118, e2109229118.

Septfons, A., Leparc-Goffart, I., Couturier, E., Franke, F., Deniau, J., Balestier, A., Guinard, A., Heuzé, G., Liebert, A.H., Mailles, A., Ndong, J.R., Poujol, I., Raguet, S., Rousseau, C., Saidouni-Oulebsir, A., Six, C., Subiros, M., Servas, V., Terrien, E., Tillaut, H., Viriot, D., Watrin, M., Wyndels, K., the Zika Surveillance Working Group, 2016. Travel-associated and autochthonous Zika virus infection in mainland France, 1 January to 15 July 2016. Eurosurveillance 21, 30315.

Subramanian, R., He, Q., Pascual, M., 2021. Quantifying asymptomatic infection and transmission of COVID-19 in New York City using observed cases, serology, and testing capacity. Proceedings of the National Academy of Sciences 118, e2019716118.

Sun, K., Wang, W., Gao, L., Wang, Y., Luo, K., Ren, L., Zhan, Z., Chen, X., Zhao, S., Huang, Y., Sun, Q., Liu, Z., Litvinova, M., Vespignani, A., Ajelli, M., Viboud, C., Yu, H., 2021. Transmission heterogeneities, kinetics, and controllability of SARS-CoV-2. Science. 10.1126/science.abe2424

Tay, J.H., Kocher, A., Duchene, S., 2024. Assessing the effect of model specification and prior sensitivity on Bayesian tests of temporal signal. PLOS Computational Biology 20, e1012371.

Tsui, J.L.-H., McCrone, J.T., Lambert, B., Bajaj, S., Inward, R.P.D., Bosetti, P., Pena, R.E., Tegally, H., Hill, V., Zarebski, A.E., Peacock, T.P., Liu, L., Wu, N., Davis, M., Bogoch, I.I., Khan, K., Kall, M., Abdul Aziz, N.I.B., Colquhoun, R., O’Toole, Á., Jackson, B., Dasgupta, A., Wilkinson, E., de Oliveira, T., COVID-19 Genomics UK (COG-UK) consortium¶, Connor, T.R., Loman, N.J., Colizza, V., Fraser, C., Volz, E., Ji, X., Gutierrez, B., Chand, M., Dellicour, S., Cauchemez, S., Raghwani, J., Suchard, M.A., Lemey, P., Rambaut, A., Pybus, O.G., Kraemer, M.U.G., 2023. Genomic assessment of invasion dynamics of SARS-CoV-2 Omicron BA.1. Science 381, 336–343.

Vaughan, T.G., Kühnert, D., Popinga, A., Welch, D., Drummond, A.J., 2014. Efficient Bayesian inference under the structured coalescent. Bioinformatics 30, 2272–2279.

Vazquez-Prokopec, G.M., Montgomery, B.L., Horne, P., Clennon, J.A., Ritchie, S.A., 2017. Combining contact tracing with targeted indoor residual spraying significantly reduces dengue transmission. Science Advances. 10.1126/sciadv.1602024

Viboud, C., Bjørnstad, O.N., Smith, D.L., Simonsen, L., Miller, M.A., Grenfell, B.T., 2006. Synchrony, Waves, and Spatial Hierarchies in the Spread of Influenza. Science. 10.1126/science.1125237

Volkov, I., Banavar, J.R., Hubbell, S.P., Maritan, A., 2003. Neutral theory and relative species abundance in ecology. Nature 424, 1035–1037.

White, F.M., 2000. Fluid Mechanics. McGraw-Hill Science, Engineering & Mathematics.

Yan, X.-Y., Wang, W.-X., Gao, Z.-Y., Lai, Y.-C., 2017. Universal model of individual and population mobility on diverse spatial scales. Nature Communications 8, 1–9.

