## Supplementary Information for "Transmission lineage dynamics and the detection of viral importation in emerging epidemics"

**SI. 1: Selection criteria for relevant studies published during the COVID-19 pandemic**
As described in Section 3 in the main text, we identified and selected peer-reviewed studies estimating the number of SARS-CoV-2 introductions and the intensity of viral importation at specific locations using phylogenetic and/or phylogeographic approaches. Relevant studies satisfying at least one of the following criteria were included:

i) The study provided estimates of the sampling proportion (number of viral genomes included per confirmed case) during the relevant study period.
ii) The study provided the number of viral genomes that were included in the analysis and estimates of the total number of confirmed cases during the relevant study period, thereby allowing the sampling proportion to be estimated.
iii) The study provided the number of viral genomes that were included in the analysis, and the total number of confirmed cases during the relevant study period could be estimated either from data provided by the authors or from external public repositories, thereby allowing the sampling proportion to be estimated.

For studies investigating the importation of specific SARS-CoV-2 variants, we used relative lineage frequencies (either provided by the study or downloaded from public repositories) to estimate variant-specific reported/confirmed case numbers. For studies considering viral movements among multiple locations, the average sampling proportion weighted by the number of cases from each location is calculated. It is important to note that these sampling proportions are intended as only rough estimates of the proportion of infections that were sampled, given the substantial variability in reporting practices, as well as changes in case definitions and the proportion of asymptomatic infections over the course of the pandemic.

**SI. 2: Derivation of an analytical expression describing the time evolution of lineage size distribution assuming local exponential growth**
As described in Section 4 in the main text, we model the growth of local transmission lineages as particle movement in a one-dimensional continuous lineage size-space. With the definition of a transmission lineage as outlined in Section 2 in the main text, the time evolution of the density of these particles in size-space can be considered as solutions to the continuity equation

$$\frac{\partial n}{\partial t} + \frac{\partial}{\partial l}\left(n\frac{dl}{dt}\right) = M(t)\delta(l-1) \qquad \text{(S1)}$$

with the boundary condition $n(l,0) = 0$ for all $l \geq 1$, where $n = n(l)$ is the particle density at $l$, $M(t)$ is the true importation rate at time $t$, and $dl/dt$ is the instantaneous lineage growth rate which is a function of both time and lineage size.

Assuming that the local infected population is undergoing exponential growth (e.g., during the early stages of an outbreak), such that $dl/dt = rl$ where $r$ is a positive constant, Eq. S1 then becomes

$$\frac{\partial n}{\partial t} + \frac{\partial n}{\partial l} rl + nr = M(t)\delta(l-1) \qquad \text{(S2)}$$

which is a first-order PDE with a source term (right hand side).

Applying change of variables with $\epsilon = t$ and $\nu = \ln l - rt$, we obtain

$$\frac{\partial n}{\partial \epsilon} + nr = M(\epsilon)\delta(\nu + r\epsilon) \qquad \text{(S3)}$$

where we have also used the transformation $\delta(g(x)) = \delta(x - x_0)/|g'(x_0)|$, if $g(x)$ has a real root at $x = x_0$.

Eq. S3 can then be solved using an integrating factor to give the general solution

$$n(\epsilon,\nu) = e^{-r\epsilon} \int M(\epsilon')\delta(\nu + r\epsilon')e^{r\epsilon'} d\epsilon' + \Phi(\nu) \qquad \text{(S4)}$$

with $\Phi(v)$ being an arbitrary function of $v$.

Applying the boundary condition that $n(l, t = 0) = 0$ for all $l \geq 1$, and since $v(l, t = 0) = \ln l$, from Eq. S4 we obtain

$$\Phi(\ln l) = -M(-\ln l/r)H(\ln l/r)/l \tag{S5}$$

where $H(x)$ is the Heaviside function. If we further let $u$ =ln $l$, and given that $H(u/r) = H(u)$, we get

$$\Phi(u) = -M(-u/r)H(u)e^{-u} \tag{S6}$$

Substituting this back into the general solution (Eq. S4) and expanding the right hand side gives

$$n(\epsilon, v) = e^{-(r\epsilon+v)}M(-v/r)[H(r\epsilon + v) - H(v)] \tag{S7}$$

Finally, performing change of variables again from $\epsilon$ and $v$ back to $t$ and $l$ gives the solution

$$n(l, t) = \frac{M(t - \ln l/r)}{l}[H(l - 1) - H(l - e^{rt})] \tag{S8}$$

The second term in final solution is commonly known as a boxcar function with the general form $H(x - a) - H(x - b)$, where $a$ and $b$ are constants representing the limits of the interval over which the function gives a value of 1, and 0 otherwise. The corresponding interval of the boxcar function in Eq. S8 is $[1, e^{rt}]$, with 1 being the minimum size that a lineage can have by definition, and $e^{rt}$ being the maximum lineage size attainable up to time $t$ assuming exponential growth at rate $r$.

**SI. 3: Derivation of an analytical expression describing the time evolution of lineage size distribution assuming local logistic growth**

Following from SI. 2, if we now assume that the local infected population is undergoing logistic growth instead such that $dl/dt = rl(1 - l/K)$, where $r$ is again a positive constant and represents the initial growth rate when $l$ is small, and $K$ is the maximum lineage size attainable (also known as the carrying capacity), Eq. S1 then becomes

$$\frac{\partial n}{\partial t} + \frac{\partial n}{\partial l}rl\left(1 - \frac{l}{K}\right) + nr\left(1 - \frac{2l}{K}\right) = M(t)\delta(l - 1) \tag{S9}$$

Applying change of variables with $\epsilon = t$ and $v = \ln[l/(K - l)]$, we obtain

$$\frac{\partial n}{\partial \epsilon} + \frac{\partial n}{\partial v}r - nr\tanh\left(\frac{v}{2}\right) = M(\epsilon)\left[\frac{K}{K - 1}\right]\delta[v + \ln(K - 1)] \tag{S10}$$

where we have again used the transformation $\delta(g(x)) = \delta(x - x_0)/|g'(x_0)|$, giving the factor $K/(K - 1)$ in the right hand side of the equation.

It can be shown that any first-order PDE of the form

$$\frac{\partial w}{\partial x} + \frac{\partial w}{\partial y}a = f(x, y)w + g(x, y) \tag{S11}$$

has the general solution

$$w(x, y) = F(x, u)\left[\Phi(u) + \int \frac{g(x, u + ax)}{F(x, u)dx}\right] \tag{S12}$$

with

$$F(x, u) = \exp\left[\int f(x, u + ax)dx\right] \tag{S13}$$

where $u = y - ax$ and $\Phi(u)$ is an arbitrary function of the parameter $u$.

If we compare Eq. S10 with Eq. S11, it is straightforward to see that the above known result implies the following general solution to our first-order PDE,

$$n(\epsilon, u) = F(\epsilon, u)\left[\Phi(u) + \int M(\epsilon')[K/(K-1)]\frac{\delta[u + r\epsilon' + \ln(K-1)]}{F(\epsilon', u)d\epsilon'}\right] \tag{S14}$$

where

$$F(\epsilon, u) = \exp\left[\int r\tanh[(u + r\epsilon')/2]\,d\epsilon'\right] = A\cosh^2[(u + r\epsilon)/2] \tag{S15}$$

with $A$ being a constant of integration.

Applying the boundary condition that $n(l, t = 0) = 0$ for all $l \geq 1$, and since $u(\nu, \epsilon = 0) = \nu = \ln[l/(K - l)]$, from Eq. S14 we obtain

$$\Phi(q) = -\frac{M[-(1/r)[q + \ln(K-1)]]H[q + \ln(K-1)]}{AK/4} \tag{S16}$$

where we have let $q = \ln[l/(K - l)]$.

Substituting this back into the general solution (Eq. S12) and expanding the right hand side gives

$$n(\epsilon, u) = \frac{\cosh^2\left[\frac{u + r\epsilon}{2}\right]\left[H[u + r\epsilon + \ln(K-1)] - H(u + \ln(K-1))\right]}{K/4}M\left[-\frac{[u + \ln(K-1)]}{r}\right] \tag{S17}$$

Finally, performing change of variables again from $\epsilon$ and $\nu$ back to $t$ and $l$ gives the solution

$$n(l, t) = \frac{K}{l(K-l)}M\left[t - \frac{ln\left[\frac{l(K-1)}{K-l}\right]}{r}\right]\left[H(l-1) - H\left[l - \frac{Ke^{rt}}{[(K-1) + e^{rt}]}\right]\right] \tag{S18}$$

Note that we again have a boxcar function as the second term in our final solution, with the corresponding interval (over which the boxcar function takes a value of 1) being $[1, Ke^{rt}/[(K-1) + e^{rt}]$. By solving $dl/dt = rl(1 - l/K)$ with the boundary condition $l(t = 0) = 1$, it is easy to show that the maximum lineage size attainable at time $t$ is given by $Ke^{rt}/[(K-1) + e^{rt}]$, i.e. the right-limit of the boxcar function, as expected.

**SI. 4: A simple deterministic model of viral importation and local lineage growth**

In this simple model, we assume that the population at the recipient location is completely susceptible initially at $t = 0$. At each subsequent time step $t$, $M(t)$ infectious travellers arrive per day from the source location, with each traveller introducing a local transmission lineage of size 1 upon arrival. Once introduced, each lineage grows deterministically assuming exponential growth at rate $r$. We also assume that there is no recovery of infected individuals, such that the size of a local lineage at any given time $t$ is given by $l(t) = e^{r(t-t_0)}$, where $t_0$ is the time when the lineage was introduced.

Once a predefined amount of time T (referred to as the time of observation) has elapsed since the first viral importation event at $t = 0$, we simulate a sampling process by randomly selecting a proportion $s$ of infectious individuals at the time of observation (with equal probability regardless of their infection time). Assuming that any uncertainties and biases associated with the phylogenetic tree estimation and phylogeographic reconstruction are negligible, a local transmission lineage is considered detected if at least one of its associated members is sampled. The inferred time of importation of each detected lineage corresponds to the time at which the associated infectious traveller entered the local population. To account for the stochasticity in the sampling process, we repeat the random selection of infected individuals 50 times for each sampling proportion $s$ and time of observation T.

**SI. 5: A stochastic agent-based model of viral importation and local lineage growth**
Here we construct a stochastic agent-based model with the following key assumptions:

1) The local population consists of N individuals initially.
2) At any given time $t$, each individual can be in one of three possible states: susceptible (S), infectious (I), or recovered (R).
3) The population is well-mixed, i.e. each individual has an equal probability of coming into contact with any other individual, regardless of their infection status.

At the start (at $t = 0$), we assume that the local population is initially in a fully susceptible state with no infected individuals. At each subsequent time step $t$, we simulate the following processes:

i) **Viral importation:** $M(t)$ infectious travellers are introduced into the local population.
ii) **Contact:** Each infected individual (including infectious travellers) comes into contact with $\kappa$ individuals randomly selected from the local population, with equal selection probability regardless of their infection status.
iii) **Transmission:** Each susceptible individual who comes into contact with an infected individual becomes infected with probability $\beta$.
iv) **Recovery:** Each infected individual recovers with probability $\gamma$.

Importantly, we keep track of who-infected-whom in each transmission event at each time step; this allows us to attribute each local infection to a specific local transmission lineage resulting from a single arriving infectious traveller.

Once a predefined amount of time T (referred to as the time of observation) has elapsed since the first viral importation event at $t = 0$, we again simulate a sampling process by randomly selecting a proportion $s$ of individuals who are either still infectious (I) or have recovered (R) at the time of observation (with equal probability regardless of their infection time). Assuming that any uncertainties and biases associated with the phylogenetic tree estimation and phylogeographic reconstruction are negligible, a local transmission lineage is considered detected if at least one of its members is sampled. The inferred time of importation of each detected lineage corresponds to the time at which the associated infectious traveller entered the local population. To account for the stochasticity in the sampling process, we repeat the random selection of infected individuals 50 times for each sampling proportion $s$ and time of observation T.

The values for parameters $\beta$ (transmission probability given contact), $\gamma$ (recovery probability per unit time), and $\kappa$ (number of contacts per individual per unit time) are specified based on estimates consistent with or similar to those observed during the COVID-19 pandemic. Details of these parameter values can be found in Table S2, along with references to studies from which these estimates were extracted.

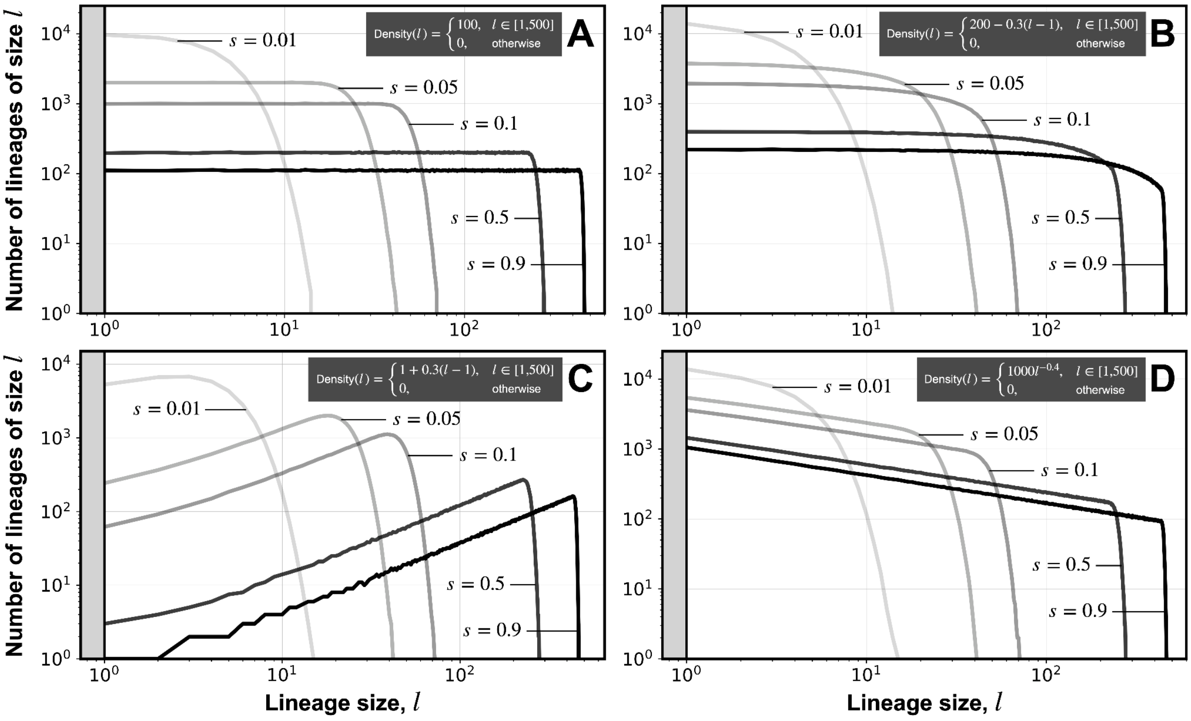


***Figure S1. Observed lineage size distribution at different sampling proportions on a log-log scale.*** *Each panel shows results from experiments considering a different true lineage size distribution (A: a uniform distribution; B: a linear increase in density with lineage size; C: a linear decrease in density with lineage size; D: a power-law distribution with a negative exponent); the corresponding density function is shown at the top of each panel. All distributions are truncated at maximum lineage size* $l = 500$*. Solid lines represent the observed lineage size distribution at varying sampling proportions (*$s = 0.01, 0.05, 0.1, 0.5, 0.9$*), as indicated by their opacity (lower opacity corresponds to a lower sampling proportion).*

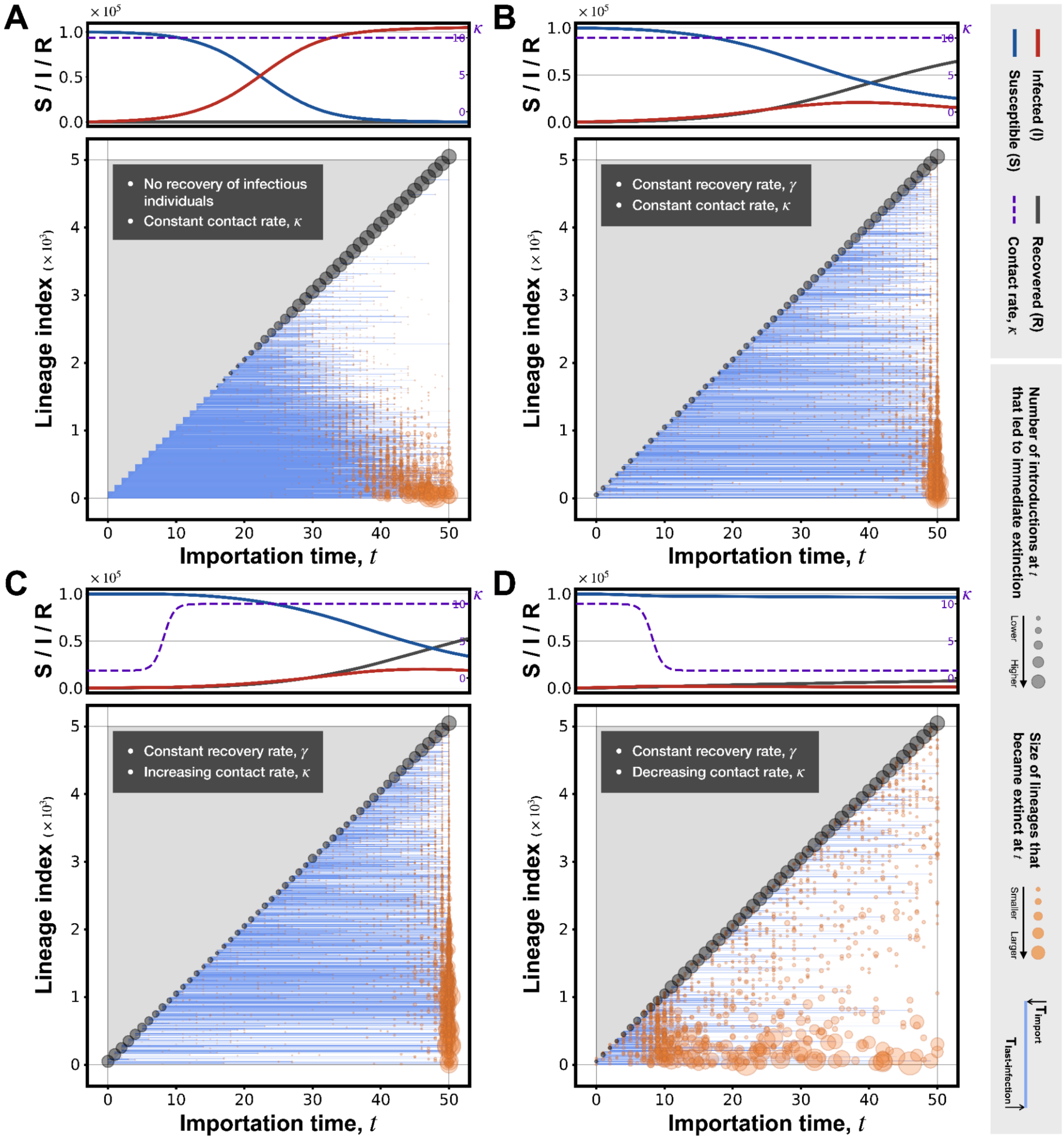


***Figure S2. Impact of local transmission dynamics on lineage growth and stochastic extinction of lineages.*** *Each panel shows results from a single stochastic agent-based simulation up to time T=50 assuming a constant importation rate ($M(t) = M_0 = 100$ per day) and a constant transmission probability ($\beta = 0.25$ per contact), but with either recovery (B, C, and D; at rate $\gamma = 1$) or without recovery (A) of infectious individuals, and either a constant contact rate (A and B; at rate $\kappa = 10$) or a time-varying contact rate (A: an increasing contact rate, from $\kappa = 1$ to $\kappa = 10$; B: a decreasing contact rate, from $\kappa = 10$ to $\kappa = 1$). The plot at the top of each panel shows the simulated epidemic dynamics, i.e. the number of infected (I; red solid line), susceptible (S; blue solid line) and recovered (R; black solid line) individuals over time. The plot at the bottom shows the time of importation and time of extinction (time of last infection) of each local transmission lineage, as represented by the start- and end-position of a horizontal blue line. Transmission lineages (horizontal blue lines) are positioned along the y-axis according to their order of importation. Extinction events are marked by orange circles positioned along the x-axis according to the time of last infection, with radius indicating the size of the lineage at extinction (i.e. total number of infected individuals since introduction). Viral introductions resulting in immediate extinction and therefore local transmission lineages of size $l = 1$ are marked by black circles, with radius indicating the number of such events at a given time $t$.*

**Table S1. Key statistics from previous studies estimating number of viral introductions and importation intensity using phylogeography during COVID-19 pandemic.** For studies in which phylogeographic analysis was performed for multiple locations, the average sampling proportion (weighted by the estimated number for reported/confirmed cases during study period) is estimated and reported; sampling proportions estimated using numbers of reported/confirmed cases taken from external data from public repositories (as opposed to from the relevant studies themselves) are indicated by an asterisk.

| ID | Title (DOI) | No. of genomes included | Estimated sampling proportion | Relevant details and/or external data used |
|---|---|---|---|---|
| 1 | A small number of early introductions seeded widespread transmission of SARS-CoV-2 in Québec, Canada (https://doi.org/10.1186/s13073-021-00986-9) (Murall et al., 2021) | 2,921 | 0.057 | N/A |
| 2 | Phylogenetic analysis of SARS-CoV-2 in Boston highlights the impact of superspreading events (https://doi.org/10.1126/science.abe3261) (Lemieux et al., 2021) | 772 | 0.012 * | Case data downloaded from https://github.com/nytimes/covid-19-data (accessed on 23 December 2024) was used to calculate cumulative number of confirmed cases in Massachusetts state between 1 March and 1 May 2020 |
| 3 | Genomic epidemiology reveals multiple introductions of SARS-CoV-2 from mainland Europe into Scotland (https://doi.org/10.1038/s41564-020-00838-z) (da Silva Filipe et al., 2021) | 1,314 | 0.49 | N/A |
| 4 | Establishment and lineage dynamics of the SARS-CoV-2 epidemic in the UK (https://doi.org/10.1126/science.abf2946) (du Plessis et al., 2021) | 26,181 | 0.093 | N/A |
| 5 | Spatiotemporal invasion dynamics of SARS-CoV-2 lineage B.1.1.7 emergence (https://doi.org/10.1126/science.abj0113) (Kraemer et al., 2021) | 17,716 | 0.038 | N/A |
| 6 | Context-specific emergence and growth of the SARS-CoV-2 Delta variant (https://doi.org/10.1038/s41586-022-05200-3) (McCrone et al., 2022) | 52,992 | 0.50 * | Total number of Delta cases in England between 12 March and 15 June 2021 was estimated using confirmed case data downloaded from OWID (https://ourworldindata.org/grapher/uk-daily-new-covid-cases?time=earliest..latest&country=~England; accessed on 12 October 2024) and lineage frequency data downloaded from https://covid19.sanger.ac.uk/lineages/raw (accessed on 13 October 2024). |
| 7 | Genomic epidemiology of the first two waves of SARS-CoV-2 in Canada (https://doi.org/10.7554/eLife.73896) (McLaughlin et al., 2022) | 27,552 | 0.045 | N/A |
| 8 | Genomic epidemiology of SARS-CoV-2 under an elimination strategy in Hong Kong (https://doi.org/10.1038/s41467-022-28420-7) (Gu et al., 2022) | 1,899 | 0.19 | Total number of confirmed cases in Hong Kong during the study period was taken from Supplementary Information provided by the authors. |
| 9 | Genomic epidemiology of the SARS-CoV-2 epidemic in Brazil | 17,135 | 0.00091 * | Total number of confirmed cases in Brazil and Paraguay |

| | | | | |
|---|---|---|---|---|
| | (https://doi.org/10.1038/s41564-022-01191-z) (Giovanetti et al., 2022) | | | up to 30 June 2021 was calculated using data downloaded from OWID (https://ourworldindata.org/coronavirus#coronavirus-country-profiles; accessed on 6 October 2024). |
| **10** | Phylogenetic estimates of SARS-CoV-2 introductions into Washington State (https://doi.org/10.1016/j.lana.2021.100018) (Tordoff et al., 2021) | 4,918 | 0.060 | N/A |
| **11** | The first wave of the COVID-19 epidemic in Spain was associated with early introductions and fast spread of a dominating genetic variant (https://doi.org/10.1038/s41588-021-00936-6) (López et al., 2021) | 2,170 | 0.0085 * | Total number of confirmed cases in Spain between 25 Feb and 22 June 2020 was calculated using data downloaded from OWID (https://ourworldindata.org/coronavirus#coronavirus-country-profiles; accessed on 23 October 2024). |
| **12** | Genomic epidemiology reveals transmission patterns and dynamics of SARS-CoV-2 in Aotearoa New Zealand (https://doi.org/10.1038/s41467-020-20235-8) (Geoghegan et al., 2020) | 649 | 0.56 | N/A |
| **13** | Genomic surveillance of SARS-CoV-2 in Puerto Rico enabled early detection and tracking of variants (https://doi.org/10.1038/s43856-022-00168-7) (Santiago et al., 2022) | 753 | 0.0040 * | Total number of confirmed cases in Puerto Rico between 23 March 2020 and 30 September 2021 was calculated using data downloaded from OWID (https://ourworldindata.org/coronavirus#coronavirus-country-profiles; accessed on 3 December 2024). |
| **14** | SARS-CoV-2 introductions and early dynamics of the epidemic in Portugal (https://doi.org/10.1038/s43856-022-00072-0) (Borges et al., 2022) | 1,275 | 0.16 | N/A |
| **15** | Genomic epidemiology of SARS-CoV-2 variants during the first two years of the pandemic in Colombia (https://doi.org/10.1038/s43856-023-00328-3) (Jiménez-Silva et al., 2023) | 1,670 | 0.00028 * | Total number of confirmed cases in Colombia up to February 2022 was calculated using data downloaded from OWID (https://ourworldindata.org/coronavirus#coronavirus-country-profiles; accessed 5 December 2024). |
| **16** | Genomic epidemiology of SARS-CoV-2 transmission lineages in Ecuador (https://doi.org/10.1093/ve/veab051) (Gutierrez et al., 2021) | 160 | 0.0008 | Total number of confirmed cases in Ecuador during the study period was taken from Supplementary Information provided by the authors. |
| **17** | Genomic epidemiology reveals the reduction of the introduction and spread of SARS-CoV-2 after implementing control strategies in Republic of Korea, 2020 (https://doi.org/10.1093/ve/veab077) (Kwon et al., 2021) | 2,065 | 0.034 | N/A |
| **18** | Epidemiological dynamics of SARS-CoV-2 VOC Gamma in Rio de Janeiro, Brazil (https://doi.org/10.1093/ve/veab087) (Moreira et al., 2021) | 113 | 0.00043 | N/A |

| | | | | |
|---|---|---|---|---|
| **19** | Sixteen novel lineages of SARS-CoV-2 in South Africa (https://doi.org/10.1038/s41591-021-01255-3) (Tegally et al., 2021) | 1,365 | 0.0022 * | Total number of confirmed cases in South Africa between 6 March and 26 August 2022 was calculated using data downloaded from OWID (https://ourworldindata.org/coronavirus#coronavirus-country-profiles; accessed on 11 November 2024). Note also that the final number of South African sequences after quality control was used in calculating the sampling proportion. |
| **20** | A year of genomic surveillance reveals how the SARS-CoV-2 pandemic unfolded in Africa (https://doi.org/10.1126/science.abj4336) (Wilkinson et al., 2021) | 8,746 | 0.002 * | Total number of confirmed cases in Africa up to 31 March 2021 was calculated using data downloaded from OWID (https://ourworldindata.org/coronavirus#coronavirus-country-profiles; accessed on 15 November 2024). |
| **21** | Evolution and epidemic spread of SARS-CoV-2 in Brazil (https://doi.org/10.1126/science.abd2161) (Candido et al., 2020) | 490 | 0.005 | N/A |
| **22** | Genomic epidemiology of SARS-CoV-2 in Guangdong Province, China (https://doi.org/10.1016/j.cell.2020.04.023) (Lu et al., 2020) | 70 | 0.05 | Total number of confirmed cases in Guangdong up to 19 March 2020 (n=1,388; including imported cases), as reported by the authors in the study, was used in calculating the sampling proportion. |
| **23** | Tracking the COVID-19 pandemic in Australia using genomics (https://doi.org/10.1038/s41467-020-18314-x) (Seemann et al., 2020) | 903 | 0.68 | The number of confirmed cases in Australia between 6 January and 14 April 2020 (n=1,333), as reported by the authors in the study, was used in calculating the sampling proportion. |
| **24** | Genomic epidemiology of the early stages of the SARS-CoV-2 outbreak in Russia (https://doi.org/10.1038/s41467-020-20880-z) (Komissarov et al., 2021) | 211 | 0.0034 | The number of confirmed cases in Russia between 11 March and 23 April 2020 (n=62,745), as reported by the authors in the study, was used in calculating the sampling proportion. |
| **25** | Genomic epidemiology of SARS-CoV-2 during the first four waves in Mozambique (https://doi.org/10.1371/journal.pgph.0001593) (Ismael et al., 2023) | 1,142 | 0.0051 | Total number of confirmed cases in Mozambique up to 22 April 2022 was calculated using data downloaded from OWID (https://ourworldindata.org/coronavirus#coronavirus-country-profiles; accessed on 20 December 2024). |
| **26** | Tracking SARS-CoV-2 introductions in Mozambique using pandemic-scale phylogenies: a retrospective observational study (https://doi.org/10.1016/S2214-109X(23)00169-9) (Martínez-Martínez et al., 2023) | 932 | 0.0067 * | Although Mozambican sequences were extracted from the GISAID database up to 11 January 2022, the study considered only the Beta and Delta variants which were the dominant lineages up to end of November 2021 (before they were overtaken by the Omicron variant) (Ismael et |

| | | | | |
|---|---|---|---|---|
| | | | | al., 2023). To calculate the sampling proportion, we have therefore used the total number of confirmed cases in Mozambique between 1 November 2020 and 30 November 2021 downloaded from OWID (https://ourworldindata.org/coronavirus#coronavirus-country-profiles; accessed on 5 January 2025), with the assumption that the proportion of confirmed cases that can be attributed to variants other than the Beta and Delta variant is negligible. |
| **27** | Tracing the international arrivals of SARS-CoV-2 Omicron variants after Aotearoa New Zealand reopened its border (https://doi.org/10.1038/s41467-022-34186-9) (Douglas et al., 2022) | 2,000 | 0.0016 * | SARS-CoV-2 transmission during the study period was dominated by the Omicron and Delta variants. To estimate the number of confirmed cases that can be attributed to Omicron, we used the total number of confirmed cases in New Zealand between 8 November 2021 and 15 June 2022 downloaded from OWID (https://ourworldindata.org/coronavirus#coronavirus-country-profiles; accessed on 15 January 2025). The number of cases attributable to the Delta variant was estimated using lineage frequency data downloaded from https://github.com/ESR-NZ/nz-sars-cov2-variants/tree/main/data (accessed on 15 January 2025); this value was then subtracted from the total number of confirmed cases during the study period to obtain the number of relevant Omicron cases. |
| **28** | Evolutionary and spatiotemporal analyses reveal multiple introductions and cryptic transmission of SARS-CoV-2 VOC/VOI in Malta (https://doi.org/10.1128/spectrum.01539-23) (Trovao et al., 2023) | 666 | 0.010 * | Total number of confirmed cases in Malta up to 25 January 2022 was calculated using data downloaded from OWID (https://ourworldindata.org/coronavirus#coronavirus-country-profiles; accessed on 12 January 2025). |
| **29** | Genomic epidemiology of early SARS-CoV-2 transmission dynamics, Gujarat, India (https://doi.org/10.3201/eid2804.212053) (Raghwani et al., 2022) | 434 | 0.0071 * | Total number of confirmed cases in Gujarat, India between 1 April and 31 July 2020 was calculated using data downloaded from https://github.com/covid19india/api (accessed on 16 January 2025).. |
| **30** | Tracking the introduction and spread of SARS-CoV-2 in coastal Kenya (https://doi.org/10.1038/s41467-021-25137-x) (Githinji et al., 2021) | 311 | 0.16 | The number of confirmed cases across the coastal counties by 31 July 2020 (n=1,997), as reported by the authors in the study, was used |

| | | | | |
|---|---|---|---|---|
| | | | | in calculating the sampling proportion. |
| **31** | A single early introduction governed viral diversity in the second wave of SARS-CoV-2 epidemic in Hungary (https://doi.org/10.1093/ve/veac069) (Ari et al., 2022) | 352 | 0.00087 | Total number of confirmed cases in Hungary between 29 April 2020 and 23 February 2021 was calculated using data downloaded from OWID (https://ourworldindata.org/coronavirus#coronavirus-country-profiles; accessed on 20 December 2024). |
| **32** | Importation of Alpha and Delta variants during the SARS-CoV-2 epidemic in Switzerland: phylogenetic analysis and intervention scenarios (https://doi.org/10.1371/journal.ppat.1011553) (Reichmuth et al., 2023) | 13,198 | 0.15 * | Total number of confirmed cases that can be attributed to the Alpha and Delta variants in Switzerland prior to 31 March 2021 and 31 July 2021, respectively, were estimated using surveillance data from Federal Office of Public Health (using an R script provided by the authors in the Supplementary Materials, https://github.com/ISPMBern/voc_imports_ch/blob/main/R/imports_100_swissepidemic.R). |
| **33** | Introduction and transmission of SARS-CoV-2 lineage B.1.1.7, Alpha variant, in Denmark (https://doi.org/10.1186/s13073-022-01045-7) (Michaelsen et al., 2022) | 60,178 | 0.34 | N/A |
| **34** | Genomic epidemiology of the first epidemic wave of severe acute respiratory syndrome coronavirus 2 (SARS-CoV-2) in Palestine (https://doi.org/10.1099/mgen.0.000584) (Qutob et al., 2021) | 69 | 0.0030 * | Total number of confirmed cases in Palestine between 4 March and19 August 2020 was calculated using data downloaded from OWID (https://ourworldindata.org/coronavirus#coronavirus-country-profiles; accessed on 10 October 2024). |
| **35** | Genomic reconstruction of the SARS-CoV-2 epidemic in England (https://doi.org/10.1038/s41586-021-04069-y) (Vöhringer et al., 2021) | 281,178 | 0.072 | N/A |
| **36** | Genomic epidemiology of the first wave of SARS-CoV-2 in Italy (https://doi.org/10.3390/v12121438) (Di Giallonardo et al., 2020) | 651 | 0.0027 * | Total number of confirmed cases in Italy between 29 January and 20 July 2020 was calculated using data downloaded from OWID (https://ourworldindata.org/coronavirus#coronavirus-country-profiles; accessed on 2 August 2024). |
| **37** | Multiple introductions followed by ongoing community spread of SARS-CoV-2 at one of the largest metropolitan areas of Northeast Brazil (https://doi.org/10.3390/v12121414) (Paiva et al., 2020) | 101 | 0.0065 * | Total number of confirmed cases in Pernambuco, Brazil between 12 March and 14 May 2020 (date of latest genome sample) was calculated using data downloaded from https://github.com/henriquemor/covid19-Brazil-timeseries/blob/master/transp-confirmed-new.csv (accessed on 25 October 2024). |
| **38** | Phylodynamics reveals the role of human | 793 | 0.079 | Sampling proportion is |

| | | | | |
|---|---|---|---|---|
| | travel and contact tracing in controlling the first wave of COVID-19 in four island nations (https://doi.org/10.1093/ve/veab052) (Douglas et al., 2021) | | | averaged across the four countries considered in the study, weighted by the number of confirmed cases per location. |
| **39** | Genomic assessment of invasion dynamics of SARS-CoV-2 Omicron BA.1 (https://doi.org/10.1126/science.adg6605) (Tsui et al., 2023) | 48,748 | 0.011 * | Total number of Omicron BA.1 cases in England up to 31 January 2021 was estimated using data on the number of confirmed cases with SGTF, as provided by the authors in the Supplementary Materials (https://github.com/joetsui1994/Omicron-BA.1-invasion-dynamics/blob/main/analyses/epidemiological/GOV.UK_SGTF_BA.1_daily_LTLA_20210901-20220301.csv; accessed on 29 August 2024). |
| **40** | Dispersal dynamics of SARS-CoV-2 lineages during the first epidemic wave in New York City (https://doi.org/10.1371/journal.ppat.1009571) (Dellicour et al., 2021) | 828 | 0.0045 * | Total number of confirmed cases in New York City (Bronx, Brooklyn, Manhattan, Queens, and Staten Island) up to 10 May 2020 was calculated using data downloaded from https://github.com/sdellicour/sars-cov-2_new_york/blob/master/Scripts_%26_data/NY_epidemiological_data/NY_boroughs_COVID.csv (accessed on 19 December 2024), as provided by the authors in the Supplementary Materials. |
| **41** | A phylodynamic workflow to rapidly gain insights into the dispersal history and dynamics of SARS-CoV-2 lineages (https://doi.org/10.1093/molbev/msaa284) (Dellicour et al., 2021) | 740 | 0.012 * | Total number of confirmed cases in Belgium up to 10 June 2020 was calculated using data downloaded from OWID (https://ourworldindata.org/coronavirus#coronavirus-country-profiles; accessed on 9 December 2024). |
| **42** | Regional connectivity drove bidirectional transmission of SARS-CoV-2 in the Middle East during travel restrictions (https://doi.org/10.1038/s41467-022-32536-1) (Parker et al., 2022) | 579 | 0.0011 | N/A |
| **43** | Genomic sequencing of SARS-CoV-2 in Rwanda reveals the importance of incoming travelers on lineage diversity (https://doi.org/10.1038/s41467-021-25985-7) (Butera et al., 2021) | 203 | 0.012 | N/A |
| **44** | SARS-CoV-2 genomic characterization and clinical manifestation of the COVID-19 outbreak in Uruguay (https://doi.org/10.1080/22221751.2020.1863747) (Elizondo et al., 2021) | 73 | 0.093 | N/A |
| **45** | Comparing the evolutionary dynamics of predominant SARS-CoV-2 virus lineages co-circulating in Mexico (https://doi.org/10.7554/eLife.82069) (Castelán-Sánchez et al., 2023) | 10,618 | 0.0042 * | Total number of confirmed cases that can be attributed to B.1.1.222, B.1.1.519, B.1.1.7, P.1, and B.1.617.2 in Mexico from January 2020 up to 30 November 2021 was estimated using confirmed |

| | | | | |
|---|---|---|---|---|
| | | | | case data downloaded from https://datos.covid-19.conacyt.mx/ (accessed on 5 January 2025) and lineage frequency data downloaded from https://cov-spectrum.org/explore/Mexico/AllSamples/ (accessed on 5 January 2025). |
| **46** | Variant-specific introduction and dispersal dynamics of SARS-CoV-2 in New York City - from Alpha to Omicron (https://doi.org/10.1371/journal.ppat.1011348) (Dellicour et al., 2023) | 12,093 | 0.0092 * | Total number of Alpha, Iota, Delta, and Omicron (BA.1) cases in New York City between 2020 and 2022 was estimated using daily confirmed case number and lineage frequency data downloaded from https://github.com/nychealth/coronavirus-data (accessed on 20 January 2024). The final sampling proportion is averaged across the four variants considered in the study, weighted by the number of confirmed cases per variant. |
| **47** | Regional importation and asymmetric within-country spread of SARS-CoV-2 variants of concern in the Netherlands (https://doi.org/10.7554/eLife.78770) (Han et al., 2022) | 3,596 | 0.0038 | Total number of Alpha, Beta, Gamma, and Delta cases in the Netherlands during the study period was estimated by calculating the total number of non-variant-specific cases between December 2020 and August 2021, and subtracting the number of cases that were not attributable to the Alpha variant between December 2020 and April 2021, using data downloaded from https://github.com/AMC-LAEB/nl_sars-cov-2_genomic_epi_2022/tree/main (accessed on 20 January 2025) as provided by the authors in the Supplementary Materials. In the calculation, we have assumed that SARS-CoV-2 transmission during the study period was dominated by the Alpha, Beta, Gamma, and Delta variants, with the proportion of other lineages being negligible. |
| **48** | Genomic evolution and early introductions of the SARS-CoV-2 Omicron variant in Mexico (https://doi.org/10.1093/ve/veac109) (Castelán-Sánchez et al., 2022) | 641 | 0.00034 * | Total number of Omicron cases in Mexico between 1 November 2021 and 31 March 2022 was estimated using data downloaded from OWID (https://ourworldindata.org/coronavirus#coronavirus-country-profiles; accessed on 12 January 2025), with the assumption that the number of cases attributable to other lineages was negligible during the study period. |

**Table S2. Key parameters used in the stochastic agent-based model.** Brief descriptions and references to studies from which values are taken are included where applicable.

| Parameter | Description | Value(s) |
|---|---|---|
| **Population size, $N$** | Size of local population (excluding arriving infectious travellers) | 100,000 |
| **Transmission probability, $\beta$** | Probability that a susceptible individual becomes infected upon contact with an infected individual | 0.025 per contact (fixed) (Ferretti et al., 2023; Thompson et al., 2021) |
| **Recovery probability, $\gamma$** | Probability that an infected individual recovers and becomes immune (i.e., I → R) per time step | 0.1 per day (fixed; equivalent to an average infectious duration of 10 days) (Byrne et al., 2020; Cevik et al., 2021) |
| **Contact rate, $\kappa$** | Average of contacts per individual per time step (regardless of infection status) | Contant $\kappa$: 10 per individual per day (Mossong et al., 2008) |
| | | Time-varying $\kappa$: increases from 1 to 10 per individual per day, following a sigmoidal trajectory (Mossong et al., 2008; Wong et al., 2023) |
| | | Time-varying $\kappa$: decreases from 10 to 1 per individual per day, following a sigmoidal trajectory (Mossong et al., 2008; Wong et al., 2023) |

### References

Ari, E., Vásárhelyi, B. M., Kemenesi, G., Tóth, G. E., Zana, B., Somogyi, B., Lanszki, Z., Röst, G., Jakab, F., Papp, B., & Kintses, B., 2022. A single early introduction governed viral diversity in the second wave of SARS-CoV-2 epidemic in Hungary. Virus evolution, 8(2), veac069.

Borges, V., Isidro, J., Trovão, N.S., Duarte, S., Cortes-Martins, H., Martiniano, H., Gordo, I., Leite, R., Vieira, L., Guiomar, R., Gomes, J.P., 2022. SARS-CoV-2 introductions and early dynamics of the epidemic in Portugal. Communications Medicine 2, 1–11.

Butera, Y., Mukantwari, E., Artesi, M., Umuringa, J.D., O'Toole, Á.N., Hill, V., Rooke, S., Hong, S.L., Dellicour, S., Majyambere, O., Bontems, S., Boujemla, B., Quick, J., Resende, P.C., Loman, N., Umumararungu, E., Kabanda, A., Murindahabi, M.M., Tuyisenge, P., Gashegu, M., Rwabihama, J.P., Sindayiheba, R., Gikic, D., Souopgui, J., Ndifon, W., Rutayisire, R., Gatare, S., Mpunga, T., Ngamije, D., Bours, V., Rambaut, A., Nsanzimana, S., Baele, G., Durkin, K., Mutesa, L., Rujeni, N., 2021. Genomic sequencing of SARS-CoV-2 in Rwanda reveals the importance of incoming travelers on lineage diversity. Nature Communications 12, 1–11.

Byrne, A.W., McEvoy, D., Collins, A.B., Hunt, K., Casey, M., Barber, A., Butler, F., Griffin, J., Lane, E.A., McAloon, C., O'Brien, K., Wall, P., Walsh, K.A., More, S.J., 2020. Inferred duration of infectious period of SARS-CoV-2: rapid scoping review and analysis of available evidence for asymptomatic and symptomatic COVID-19 cases. BMJ Open 10, e039856.

Candido, D. S., Claro, I. M., de Jesus, J. G., Souza, W. M., Moreira, F. R. R., Dellicour, S., Mellan, T. A., du Plessis, L., Pereira, R. H. M., Sales, F. C. S., Manuli, E. R., Thézé, J., Almeida, L., Menezes, M. T., Voloch, C. M., Fumagalli, M. J., Coletti, T. M., da Silva, C. A. M., Ramundo, M. S., Amorim, M. R., … Faria, N. R., 2020. Evolution and epidemic spread of SARS-CoV-2 in Brazil. Science (New York, N.Y.), 369(6508), 1255–1260.

Castelán-Sánchez, H. G., Martínez-Castilla, L. P., Sganzerla-Martínez, G., Torres-Flores, J., & López-Leal, G. (2022). Genome Evolution and Early Introductions of the SARS-CoV-2 Omicron Variant in Mexico. Virus evolution, 8(2), veac109.

Castelán-Sánchez, H.G., Delaye, L., Inward, R.P.D., Dellicour, S., Gutierrez, B., de la Vina, N.M., Boukadida, C., Pybus, O.G., de Anda Jáuregui, G., Guzmán, P., Flores-Garrido, M., Fontanelli, Ó., Rosales, M.H., Meneses, A., Olmedo-Alvarez, G., Herrera-Estrella, A.H., Sánchez-Flores, A., Muñoz-Medina, J.E., Comas-García, A., Gómez-Gil, B., Zárate, S., Taboada, B., López, S., Arias, C.F., Kraemer, M.U.G., Lazcano, A., Zamudio, M.E., 2023. Comparing the evolutionary dynamics of predominant SARS-CoV-2 virus lineages co-circulating in Mexico.

Cevik, M., Tate, M., Lloyd, O., Maraolo, A.E., Schafers, J., Ho, A., 2021. SARS-CoV-2, SARS-CoV, and MERS-CoV viral load dynamics, duration of viral shedding, and infectiousness: a systematic review and meta-analysis. The Lancet. Microbe 2.

da Silva Filipe, A., Shepherd, J.G., Williams, T., Hughes, J., Aranday-Cortes, E., Asamaphan, P., Ashraf, S., Balcazar, C., Brunker, K., Campbell, A., Carmichael, S., Davis, C., Dewar, R., Gallagher, M.D., Gunson, R., Hill, V., Ho, A., Jackson, B., James, E., Jesudason, N., Johnson, N., McWilliam Leitch, E.C., Li, K., MacLean, A., Mair, D., McAllister, D.A., McCrone, J.T., McDonald, S.E., McHugh, M.P., Morris, A.K., Nichols, J., Niebel, M., Nomikou, K., Orton, R.J., O'Toole, Á., Palmarini, M., Parcell, B.J., Parr, Y.A., Rambaut, A., Rooke, S., Shaaban, S., Shah, R., Singer, J.B., Smollett, K., Starinskij, I., Tong, L., Sreenu, V.B., Wastnedge, E., Holden, M.T.G., Robertson, D.L., Templeton, K., Thomson, E.C., 2020. Genomic epidemiology reveals multiple introductions of SARS-CoV-2 from mainland Europe into Scotland. Nature Microbiology 6, 112–122.

Dellicour, S., Durkin, K., Hong, S.L., Vanmechelen, B., Martí-Carreras, J., Gill, M.S., Meex, C., Bontems, S., André, E., Gilbert, M., Walker, C., Maio, N.D., Faria, N.R., Hadfield, J., Hayette, M.-P., Bours, V., Wawina-Bokalanga, T., Artesi, M., Baele, G., Maes, P., 2020. A Phylodynamic Workflow to Rapidly Gain Insights into the Dispersal History and Dynamics of SARS-CoV-2 Lineages. Mol. Biol. Evol. 38, 1608–1613.

Dellicour, S., Hong, S.L., Hill, V., Dimartino, D., Marier, C., Zappile, P., Harkins, G.W., Lemey, P., Baele, G., Duerr, R., Heguy, A., 2023. Variant-specific introduction and dispersal dynamics of SARS-CoV-2 in New York City – from Alpha to Omicron. PLOS Pathogens 19, e1011348.

Dellicour, S., Hong, S.L., Vrancken, B., Chaillon, A., Gill, M.S., Maurano, M.T., Ramaswami, S., Zappile, P., Marier, C., Harkins, G.W., Baele, G., Duerr, R., Heguy, A., 2021. Dispersal dynamics of SARS-CoV-2 lineages during the first epidemic wave in New York City. PLOS Pathogens 17, e1009571.

Di Giallonardo, F., Duchene, S., Puglia, I., Curini, V., Profeta, F., Cammà, C., Marcacci, M., Calistri, P., Holmes, E.C., Lorusso, A., 2020. Genomic Epidemiology of the First Wave of SARS-CoV-2 in Italy. Viruses 12, 1438.

Douglas, J., Mendes, F. K., Bouckaert, R., Xie, D., Jiménez-Silva, C. L., Swanepoel, C., de Ligt, J., Ren, X., Storey, M., Hadfield, J., Simpson, C. R., Geoghegan, J. L., Drummond, A. J., & Welch, D., 2021. Phylodynamics reveals the role of human travel and contact tracing in controlling the first wave of COVID-19 in four island nations. Virus evolution, 7(2), veab052.

Douglas, J., Winter, D., McNeill, A., Carr, S., Bunce, M., French, N., Hadfield, J., de Ligt, J., Welch, D., Geoghegan, J.L., 2022. Tracing the international arrivals of SARS-CoV-2 Omicron variants after Aotearoa New Zealand reopened its border. Nature Communications 13, 1–10.

du Plessis, L., McCrone, J.T., Zarebski, A.E., Hill, V., Ruis, C., Gutierrez, B., Raghwani, J., Ashworth, J., Colquhoun, R., Connor, T.R., Faria, N.R., Jackson, B., Loman, N.J., O'Toole, Á., Nicholls, S.M., Parag, K.V., Scher, E., Vasylyeva, T.I., Volz, E.M., Watts, A., Bogoch, I.I., Khan, K., COVID-19 Genomics UK (COG-UK) Consortium†, Aanensen, D.M., Kraemer, M.U.G., Rambaut, A., Pybus, O.G., 2021. Establishment and lineage dynamics of the SARS-CoV-2 epidemic in the UK. Science.

Elizondo, V., Harkins, G.W., Mabvakure, B., Smidt, S., Zappile, P., Marier, C., Maurano, M.T., Perez, V., Mazza, N., Beloso, C., Ifran, S., Fernandez, M., Santini, A., Perez, V., Estevez, V., Nin, M., Manrique, G., Perez, L., Ross, F.,

Boschi, S., Zubillaga, M.N., Balleste, R., Dellicour, S., Heguy, A., Duerr, R., 2021. SARS-CoV-2 genomic characterization and clinical manifestation of the COVID-19 outbreak in Uruguay. Emerging Microbes & Infections.

Ferretti, L., Wymant, C., Petrie, J., Tsallis, D., Kendall, M., Ledda, A., Di Lauro, F., Fowler, A., Di Francia, A., Panovska-Griffiths, J., Abeler-Dörner, L., Charalambides, M., Briers, M., Fraser, C., 2023. Digital measurement of SARS-CoV-2 transmission risk from 7 million contacts. Nature 626, 145–150.

Geoghegan, J.L., Ren, X., Storey, M., Hadfield, J., Jelley, L., Jefferies, S., Sherwood, J., Paine, S., Huang, S., Douglas, J., Mendes, F.K., Sporle, A., Baker, M.G., Murdoch, D.R., French, N., Simpson, C.R., Welch, D., Drummond, A.J., Holmes, E.C., Duchêne, S., de Ligt, J., 2020. Genomic epidemiology reveals transmission patterns and dynamics of SARS-CoV-2 in Aotearoa New Zealand. Nature Communications 11, 1–7.

Giovanetti, M., Slavov, S.N., Fonseca, V., Wilkinson, E., Tegally, H., Patané, J.S.L., Viala, V.L., San, E.J., Rodrigues, E.S., Santos, E.V., Aburjaile, F., Xavier, J., Fritsch, H., Adelino, T.E.R., Pereira, F., Leal, A., Iani, F.C. de M., de Carvalho Pereira, G., Vazquez, C., Sanabria, G.M.E., Oliveira, E.C. de, Demarchi, L., Croda, J., dos Santos Bezerra, R., Paola Oliveira de Lima, L., Martins, A.J., Renata dos Santos Barros, C., Marqueze, E.C., de Souza Todao Bernardino, J., Moretti, D.B., Brassaloti, R.A., de Lello Rocha Campos Cassano, R., Mariani, P.D.S.C., Kitajima, J.P., Santos, B., Proto-Siqueira, R., Cantarelli, V.V., Tosta, S., Nardy, V.B., Reboredo de Oliveira da Silva, L., Gómez, M.K.A., Lima, J.G., Ribeiro, A.A., Guimarães, N.R., Watanabe, L.T., Barbosa Da Silva, L., da Silva Ferreira, R., da Penha, M.P.F., Ortega, M.J., de la Fuente, A.G., Villalba, S., Torales, J., Gamarra, M.L., Aquino, C., Figueredo, G.P.M., Fava, W.S., Motta-Castro, A.R.C., Venturini, J., do Vale Leone de Oliveira, S.M., Gonçalves, C.C.M., do Carmo Debur Rossa, M., Becker, G.N., Giacomini, M.P., Marques, N.Q., Riediger, I.N., Raboni, S., Mattoso, G., Cataneo, A.D., Zanluca, C., Duarte dos Santos, C.N., Assato, P.A., Allan da Silva da Costa, F., Poleti, M.D., Lesbon, J.C.C., Mattos, E.C., Banho, C.A., Sacchetto, L., Moraes, M.M., Grotto, R.M.T., Souza-Neto, J.A., Nogueira, M.L., Fukumasu, H., Coutinho, L.L., Calado, R.T., Neto, R.M., Bispo de Filippis, A.M., Venancio da Cunha, R., Freitas, C., Peterka, C.R.L., de Fátima Rangel Fernandes, C., Navegantes, W., do Carmo Said, R.F., Campelo de A e Melo, C.F., Almiron, M., Lourenço, J., de Oliveira, T., Holmes, E.C., Haddad, R., Sampaio, S.C., Elias, M.C., Kashima, S., Junior de Alcantara, L.C., Covas, D.T., 2022. Genomic epidemiology of the SARS-CoV-2 epidemic in Brazil. Nature Microbiology 7, 1490–1500.

Githinji, G., de Laurent, Z.R., Mohammed, K.S., Omuoyo, D.O., Macharia, P.M., Morobe, J.M., Otieno, E., Kinyanjui, S.M., Agweyu, A., Maitha, E., Kitole, B., Suleiman, T., Mwakinangu, M., Nyambu, J., Otieno, J., Salim, B., Kasera, K., Kiiru, J., Aman, R., Barasa, E., Warimwe, G., Bejon, P., Tsofa, B., Ochola-Oyier, L.I., Nokes, D.J., Agoti, C.N., 2021. Tracking the introduction and spread of SARS-CoV-2 in coastal Kenya. Nature Communications 12, 1–10.

Gu, H., Xie, R., Adam, D.C., Tsui, J.L.-H., Chu, D.K., Chang, L.D.J., Cheuk, S.S.Y., Gurung, S., Krishnan, P., Ng, D.Y.M., Liu, G.Y.Z., Wan, C.K.C., Cheng, S.S.M., Edwards, K.M., Leung, K.S.M., Wu, J.T., Tsang, D.N.C., Leung, G.M., Cowling, B.J., Peiris, M., Lam, T.T.Y., Dhanasekaran, V., Poon, L.L.M., 2022. Genomic epidemiology of SARS-CoV-2 under an elimination strategy in Hong Kong. Nature Communications 13, 1–10.

Gutierrez, B., Márquez, S., Prado-Vivar, B., Becerra-Wong, M., Guadalupe, J.J., Candido, D.D.S., Fernandez-Cadena, J.C., Morey-Leon, G., Armas-Gonzalez, R., Andrade-Molina, D.M., Bruno, A., De Mora, D., Olmedo, M., Portugal, D., Gonzalez, M., Orlando, A., Drexler, J.F., Moreira-Soto, A., Sander, A.-L., Brünink, S., Kühne, A., Patiño, L., Carrazco-Montalvo, A., Mestanza, O., Zurita, J., Sevillano, G., Du Plessis, L., McCrone, J.T., Coloma, J., Trueba, G., Barragán, V., Rojas-Silva, P., Grunauer, M., Kraemer, M.U.G., Faria, N.R., Escalera-Zamudio, M., Pybus, O.G., Cárdenas, P., 2021. Genomic epidemiology of SARS-CoV-2 transmission lineages in Ecuador. Virus Evol 7, veab051.

Han, A.X., Kozanli, E., Koopsen, J., Vennema, H., RIVM COVID-19 molecular epidemiology group, Aarts, L., Bos, S., van den Brandt, A., van den Brink, S., Cremer, J., Freriks, K., Jaarsma, R., Schmitz, D., Then, E., van der Veer, B., Wijsman, L., Zwagemaker, F., Hajji, K., Kroneman, A., van Walle, I., Klinkenberg, D., Wallinga, J., Russell, C.A., Eggink, D., Reusken, C., 2022. Regional importation and asymmetric within-country spread of SARS-CoV-2 variants of concern in the Netherlands.

Ismael, N., van Wyk, S., Tegally, H., Giandhari, J., San, J.E., Moir, M., Pillay, S., Utpatel, C., Singh, L., Naidoo, Y., Ramphal, U., Mabunda, N., Abílio, N., Arnaldo, P., Xavier, J., Amoako, D.G., Everatt, J., Ramphal, Y., Maharaj, A., de Araujo, L., Anyaneji, U.J., Tshiabuila, D., Viegas, S., Lessells, R., Engelbrecht, S., Gudo, E., Jani, I., Niemann, S., Wilkinson, E., de Oliveira, T., 2023. Genomic epidemiology of SARS-CoV-2 during the first four waves in Mozambique. PLOS Global Public Health 3, e0001593.

Jimenez-Silva, C., Rivero, R., Douglas, J., Bouckaert, R., Villabona-Arenas, C.J., Atkins, K.E., Gastelbondo, B., Calderon, A., Guzman, C., Echeverri-De la Hoz, D., Muñoz, M., Ballesteros, N., Castañeda, S., Patiño, L.H., Ramirez, A., Luna, N., Paniz-Mondolfi, A., Serrano-Coll, H., Ramirez, J.D., Mattar, S., Drummond, A.J., 2023. Genomic epidemiology of SARS-CoV-2 variants during the first two years of the pandemic in Colombia. Communications Medicine 3, 1–12.

Komissarov, A.B., Safina, K.R., Garushyants, S.K., Fadeev, A.V., Sergeeva, M.V., Ivanova, A.A., Danilenko, D.M., Lioznov, D., Shneider, O.V., Shvyrev, N., Spirin, V., Glyzin, D., Shchur, V., Bazykin, G.A., 2021. Genomic epidemiology of the early stages of the SARS-CoV-2 outbreak in Russia. Nature Communications 12, 1–13.

Kraemer, M.U.G., Hill, V., Ruis, C., Dellicour, S., Bajaj, S., McCrone, J.T., Baele, G., Parag, K.V., Battle, A.L., Gutierrez, B., Jackson, B., Colquhoun, R., O'Toole, Á., Klein, B., Vespignani, A., COVID-19 Genomics UK (COG-UK) Consortium‡, Volz, E., Faria, N.R., Aanensen, D.M., Loman, N.J., du Plessis, L., Cauchemez, S., Rambaut, A., Scarpino, S.V., Pybus, O.G., 2021. Spatiotemporal invasion dynamics of SARS-CoV-2 lineage B.1.1.7 emergence. Science.

Kwon, J.-H., Kim, J.-M., Lee, D.-H., Park, A.K., Kim, I.-H., Kim, D.-W., Kim, J.-Y., Lim, N., Cho, K.-Y., Kim, H.M., Lee, N.-J., Woo, S., Lee, C.Y., No, J.S., Kim, J., Rhee, J., Han, M.-G., Rhie, G.-E., Yoo, C.K., Kim, E.-J., 2021. Genomic epidemiology reveals the reduction of the introduction and spread of SARS-CoV-2 after implementing control strategies in Republic of Korea, 2020. Virus Evol 7, veab077.

Lemieux, J.E., Siddle, K.J., Shaw, B.M., Loreth, C., Schaffner, S.F., Gladden-Young, A., Adams, G., Fink, T., Tomkins-Tinch, C.H., Krasilnikova, L.A., DeRuff, K.C., Rudy, M., Bauer, M.R., Lagerborg, K.A., Normandin, E., Chapman,

S.B., Reilly, S.K., Anahtar, M.N., Lin, A.E., Carter, A., Myhrvold, C., Kemball, M.E., Chaluvadi, S., Cusick, C., Flowers, K., Neumann, A., Cerrato, F., Farhat, M., Slater, D., Harris, J.B., Branda, J.A., Hooper, D., Gaeta, J.M., Baggett, T.P., O'Connell, J., Gnirke, A., Lieberman, T.D., Philippakis, A., Burns, M., Brown, C.M., Luban, J., Ryan, E.T., Turbett, S.E., LaRocque, R.C., Hanage, W.P., Gallagher, G.R., Madoff, L.C., Smole, S., Pierce, V.M., Rosenberg, E., Sabeti, P.C., Park, D.J., MacInnis, B.L., 2021. Phylogenetic analysis of SARS-CoV-2 in Boston highlights the impact of superspreading events. Science.

López, M.G., Chiner-Oms, Á., García de Viedma, D., Ruiz-Rodriguez, P., Bracho, M.A., Cancino-Muñoz, I., D'Auria, G., de Marco, G., García-González, N., Goig, G.A., Gómez-Navarro, I., Jiménez-Serrano, S., Martinez-Priego, L., Ruiz-Hueso, P., Ruiz-Roldán, L., Torres-Puente, M., Alberola, J., Albert, E., Aranzamendi Zaldumbide, M., Bea-Escudero, M.P., Boga, J.A., Bordoy, A.E., Canut-Blasco, A., Carvajal, A., Cilla Eguiluz, G., Cordón Rodríguez, M.L., Costa-Alcalde, J.J., de Toro, M., de Toro Peinado, I., del Pozo, J.L., Duchêne, S., Fernández-Pinero, J., Fuster Escrivá, B., Gimeno Cardona, C., González Galán, V., Gonzalo Jiménez, N., Hernáez Crespo, S., Herranz, M., Lepe, J.A., López-Causapé, C., López-Hontangas, J.L., Martín, V., Martró, E., Milagro Beamonte, A., Montes Ros, M., Moreno-Muñoz, R., Navarro, D., Navarro-Marí, J.M., Not, A., Oliver, A., Palop-Borrás, B., Parra Grande, M., Pedrosa-Corral, I., Pérez González, M.C., Pérez-Lago, L., Pérez-Ruiz, M., Piñeiro Vázquez, L., Rabella, N., Rezusta, A., Robles Fonseca, L., Rodríguez-Villodres, Á., Sanbonmatsu-Gámez, S., Sicilia, J., Soriano, A., Tirado Balaguer, M.D., Torres, I., Tristancho, A., Marimón, J.M., Coscolla, M., González-Candelas, F., Comas, I., 2021. The first wave of the COVID-19 epidemic in Spain was associated with early introductions and fast spread of a dominating genetic variant. Nature Genetics 53, 1405–1414.

Lu, J., du Plessis, L., Liu, Z., Hill, V., Kang, M., Lin, H., Sun, J., François, S., Kraemer, M. U. G., Faria, N. R., McCrone, J. T., Peng, J., Xiong, Q., Yuan, R., Zeng, L., Zhou, P., Liang, C., Yi, L., Liu, J., Xiao, J., … Ke, C., 2020. Genomic Epidemiology of SARS-CoV-2 in Guangdong Province, China. Cell, 181(5), 997–1003.e9.

Martínez-Martínez, F. J., Massinga, A. J., De Jesus, Á., Ernesto, R. M., Cano-Jiménez, P., Chiner-Oms, Á., Gómez-Navarro, I., Guillot-Fernández, M., Guinovart, C., Sitoe, A., Vubil, D., Bila, R., Gujamo, R., Enosse, S., Jiménez-Serrano, S., Torres-Puente, M., Comas, I., Mandomando, I., López, M. G., & Mayor, A., 2023. Tracking SARS-CoV-2 introductions in Mozambique using pandemic-scale phylogenies: a retrospective observational study. The Lancet. Global health, 11(6), e933–e941.

McCrone, J.T., Hill, V., Bajaj, S., Pena, R.E., Lambert, B.C., Inward, R., Bhatt, S., Volz, E., Ruis, C., Dellicour, S., Baele, G., Zarebski, A.E., Sadilek, A., Wu, N., Schneider, A., Ji, X., Raghwani, J., Jackson, B., Colquhoun, R., O'Toole, Á., Peacock, T.P., Twohig, K., Thelwall, S., Dabrera, G., Myers, R., Faria, N.R., Huber, C., Bogoch, I.I., Khan, K., du Plessis, L., Barrett, J.C., Aanensen, D.M., Barclay, W.S., Chand, M., Connor, T., Loman, N.J., Suchard, M.A., Pybus, O.G., Rambaut, A., Kraemer, M.U.G., 2022. Context-specific emergence and growth of the SARS-CoV-2 Delta variant. Nature 610, 154–160.

McLaughlin, A., Montoya, V., Miller, R.L., Mordecai, G.J., Canadian COVID-19 Genomics Network (CanCOGen) Consortium, Worobey, M., Poon, A.F.Y., Joy, J.B., 2022. Genomic epidemiology of the first two waves of SARS-CoV-2 in Canada.

Michaelsen, T.Y., Bennedbæk, M., Christiansen, L.E., Jørgensen, M.S.F., Møller, C.H., Sørensen, E.A., Knutsson, S., Brandt, J., Jensen, T.B.N., Chiche-Lapierre, C., Collados, E.F., Sørensen, T., Petersen, C., Le-Quy, V., Sereika, M., Hansen, F.T., Rasmussen, M., Fonager, J., Karst, S.M., Marvig, R.L., Stegger, M., Sieber, R.N., Skov, R., Legarth, R., Krause, T.G., Fomsgaard, A., Albertsen, M., 2022. Introduction and transmission of SARS-CoV-2 lineage B.1.1.7, Alpha variant, in Denmark. Genome Medicine 14, 1–13.

Moreira, F. R. R., D'arc, M., Mariani, D., Herlinger, A. L., Schiffler, F. B., Rossi, Á. D., Leitão, I. C., Miranda, T. D. S., Cosentino, M. A. C., Tôrres, M. C. P., da Costa, R. M. D. S. C., Gonçalves, C. C. A., Faffe, D. S., Galliez, R. M., Junior, O. D. C. F., Aguiar, R. S., Dos Santos, A. F. A., Voloch, C. M., Castiñeiras, T. M. P. P., & Tanuri, A., 2021. Epidemiological dynamics of SARS-CoV-2 VOC Gamma in Rio de Janeiro, Brazil. Virus evolution, 7(2), veab087.

Mossong, J., Hens, N., Jit, M., Beutels, P., Auranen, K., Mikolajczyk, R., Massari, M., Salmaso, S., Tomba, G.S., Wallinga, J., Heijne, J., Sadkowska-Todys, M., Rosinska, M., John Edmunds, W., 2008. Social Contacts and Mixing Patterns Relevant to the Spread of Infectious Diseases. PLOS Medicine 5, e74.

Murall, C.L., Fournier, E., Galvez, J.H., N'Guessan, A., Reiling, S.J., Quirion, P.-O., Naderi, S., Roy, A.-M., Chen, S.-H., Stretenowich, P., Bourgey, M., Bujold, D., Gregoire, R., Lepage, P., St-Cyr, J., Willet, P., Dion, R., Charest, H., Lathrop, M., Roger, M., Bourque, G., Ragoussis, J., Shapiro, B.J., Moreira, S., 2021. A small number of early introductions seeded widespread transmission of SARS-CoV-2 in Québec, Canada. Genome Medicine 13, 1–17.

Paiva, M.H.S., Guedes, D.R.D., Docena, C., Bezerra, M.F., Dezordi, F.Z., Machado, L.C., Krokovsky, L., Helvecio, E., da Silva, A.F., Vasconcelos, L.R.S., Rezende, A.M., da Silva, S.J.R., Sales, K.G. da S., de Sá, B.S.L.F., da Cruz, D.L., Cavalcanti, C.E., Neto, A. de M., da Silva, C.T.A., Mendes, R.P.G., da Silva, M.A.L., Gräf, T., Resende, P.C., Bello, G., Barros, M. da S., do Nascimento, W.R.C., Arcoverde, R.M.L., Bezerra, L.C.A., Brandão-Filho, S.P., Ayres, C.F.J., Wallau, G.L., 2020. Multiple Introductions Followed by Ongoing Community Spread of SARS-CoV-2 at One of the Largest Metropolitan Areas of Northeast Brazil. Viruses 12, 1414.

Parker, E., Anderson, C., Zeller, M., Tibi, A., Havens, J.L., Laroche, G., Benlarbi, M., Ariana, A., Robles-Sikisaka, R., Latif, A.A., Watts, A., Awidi, A., Jaradat, S.A., Gangavarapu, K., Ramesh, K., Kurzban, E., Matteson, N.L., Han, A.X., Hughes, L.D., McGraw, M., Spencer, E., Nicholson, L., Khan, K., Suchard, M.A., Wertheim, J.O., Wohl, S., Côté, M., Abdelnour, A., Andersen, K.G., Abu-Dayyeh, I., 2022. Regional connectivity drove bidirectional transmission of SARS-CoV-2 in the Middle East during travel restrictions. Nature Communications 13, 1–14.

Qutob, N., Salah, Z., Richard, D., Darwish, H., Sallam, H., Shtayeh, I., Najjar, O., Ruzayqat, M., Najjar, D., Balloux, F., van Dorp, L., 2021. Genomic epidemiology of the first epidemic wave of severe acute respiratory syndrome coronavirus 2 (SARS-CoV-2) in Palestine. Microbial Genomics 7, 000584.

Raghwani, J., du Plessis, L., McCrone, J. T., Hill, S. C., Parag, K. V., Thézé, J., Kumar, D., Puvar, A., Pandit, R., Pybus, O. G., Fournié, G., Joshi, M., & Joshi, C., 2022. Genomic Epidemiology of Early SARS-CoV-2 Transmission Dynamics, Gujarat, India. Emerging infectious diseases, 28(4), 751–758.

Reichmuth, M.L., Hodcroft, E.B., Althaus, C.L., 2023. Importation of Alpha and Delta variants during the SARS-CoV-2 epidemic in Switzerland: Phylogenetic analysis and intervention scenarios. PLOS Pathogens 19, e1011553.

Santiago, G.A., Flores, B., González, G.L., Charriez, K.N., Huertas, L.C., Volkman, H.R., Van Belleghem, S.M., Rivera-Amill, V., Adams, L.E., Marzán, M., Hernández, L., Cardona, I., O'Neill, E., Paz-Bailey, G., Papa, R., Muñoz-Jordan, J.L., 2022. Genomic surveillance of SARS-CoV-2 in Puerto Rico enabled early detection and tracking of variants. Communications Medicine 2, 1–11.

Seemann, T., Lane, C.R., Sherry, N.L., Duchene, S., Gonçalves da Silva, A., Caly, L., Sait, M., Ballard, S.A., Horan, K., Schultz, M.B., Hoang, T., Easton, M., Dougall, S., Stinear, T.P., Druce, J., Catton, M., Sutton, B., van Diemen, A., Alpren, C., Williamson, D.A., Howden, B.P., 2020. Tracking the COVID-19 pandemic in Australia using genomics. Nature Communications 11, 1–9.

Tegally, H., Wilkinson, E., Lessells, R.J., Giandhari, J., Pillay, S., Msomi, N., Mlisana, K., Bhiman, J.N., von Gottberg, A., Walaza, S., Fonseca, V., Allam, M., Ismail, A., Glass, A.J., Engelbrecht, S., Van Zyl, G., Preiser, W., Williamson, C., Petruccione, F., Sigal, A., Gazy, I., Hardie, D., Hsiao, N.-Y., Martin, D., York, D., Goedhals, D., San, E.J., Giovanetti, M., Lourenço, J., Alcantara, L.C.J., de Oliveira, T., 2021. Sixteen novel lineages of SARS-CoV-2 in South Africa. Nature Medicine 27, 440–446.

Thompson, H.A., Mousa, A., Dighe, A., Fu, H., Arnedo-Pena, A., Barrett, P., Bellido-Blasco, J., Bi, Q., Caputi, A., Chaw, L., De Maria, L., Hoffmann, M., Mahapure, K., Ng, K., Raghuram, J., Singh, G., Soman, B., Soriano, V., Valent, F., Vimercati, L., Wee, L.E., Wong, J., Ghani, A.C., Ferguson, N.M., 2021. Severe Acute Respiratory Syndrome Coronavirus 2 (SARS-CoV-2) Setting-specific Transmission Rates: A Systematic Review and Meta-analysis. Clin Infect Dis 73, e754–e764.

Tordoff, D. M., Greninger, A. L., Roychoudhury, P., Shrestha, L., Xie, H., Jerome, K. R., Breit, N., Huang, M.-L., Famulare, M., & Herbeck, J. T., 2021. Phylogenetic estimates of SARS-CoV-2 introductions into Washington State. The Lancet Regional Health – Americas, 1, 100018.

Trovao, N.S., Pan, V., Goel, C., Gallego-García, P., Liu, Y., Barbara, C., Borg, R., Briffa, M., Cilia, C., Grech, L., Vassallo, M., Treangen, T.J., Posada, D., Beheshti, A., Borg, J., Zahra, G., 2023. Evolutionary and spatiotemporal analyses reveal multiple introductions and cryptic transmission of SARS-CoV-2 VOC/VOI in Malta. Microbiology Spectrum.

Tsui, J. L., McCrone, J. T., Lambert, B., Bajaj, S., Inward, R. P. D., Bosetti, P., Pena, R. E., Tegally, H., Hill, V., Zarebski, A. E., Peacock, T. P., Liu, L., Wu, N., Davis, M., Bogoch, I. I., Khan, K., Kall, M., Abdul Aziz, N. I. B., Colquhoun, R., O'Toole, Á., … Kraemer, M. U. G., 2023. Genomic assessment of invasion dynamics of SARS-CoV-2 Omicron BA.1. Science (New York, N.Y.), 381(6655), 336–343.

Vöhringer, H.S., Sanderson, T., Sinnott, M., De Maio, N., Nguyen, T., Goater, R., Schwach, F., Harrison, I., Hellewell, J., Ariani, C.V., Gonçalves, S., Jackson, D.K., Johnston, I., Jung, A.W., Saint, C., Sillitoe, J., Suciu, M., Goldman, N., Panovska-Griffiths, J., Birney, E., Volz, E., Funk, S., Kwiatkowski, D., Chand, M., Martincorena, I., Barrett, J.C., Gerstung, M., 2021. Genomic reconstruction of the SARS-CoV-2 epidemic in England. Nature 600, 506–511.

Wilkinson, E., Giovanetti, M., Tegally, H., San, J. E., Lessells, R., Cuadros, D., Martin, D. P., Rasmussen, D. A., Zekri, A. N., Sangare, A. K., Ouedraogo, A. S., Sesay, A. K., Priscilla, A., Kemi, A. S., Olubusuyi, A. M., Oluwapelumi, A. O. O., Hammami, A., Amuri, A. A., Sayed, A., Ouma, A. E. O., … de Oliveira, T., 2021. A year of genomic surveillance reveals how the SARS-CoV-2 pandemic unfolded in Africa. Science (New York, N.Y.), 374(6566), 423–431.

Wong, K.L.M., Gimma, A., Coletti, P., CoMix Europe Working Group, Faes, C., Beutels, P., Hens, N., Jaeger, V.K., Karch, A., Johnson, H., Edmunds, W., Jarvis, C.I., 2023. Social contact patterns during the COVID-19 pandemic in 21 European countries – evidence from a two-year study. BMC Infectious Diseases 23, 268.
